# Distributed Statistical Analyses: A Scoping Review and Examples of Operational Frameworks Adapted to Healthcare

**DOI:** 10.1101/2023.12.21.23300389

**Authors:** Félix Camirand Lemyre, Simon Lévesque, Marie-Pier Domingue, Klaus Herrmann, Jean-François Ethier

## Abstract

Data from multiple organizations are crucial for advancing learning health systems. However, ethical, legal, and social concerns may restrict the use of standard statistical methods that rely on pooling data. Although distributed algorithms offer alternatives, they may not always be suitable for healthcare research frameworks. This paper aims to support researchers and data custodians in three ways: (1) providing a concise overview of the literature on statistical inference methods for horizontally partitioned data; (2) describing the methods applicable to generalized linear models (GLM) and assessing their underlying distributional assumptions; (3) adapting existing methods to make them fully usable in healthcare research. A scoping review methodology was employed for the literature mapping, from which methods presenting a methodological framework for GLM analyses with horizontally partitioned data were identified and assessed from the perspective of applicability in healthcare research. From the review, 41 articles were selected, and six approaches were extracted for conducting standard GLM-based statistical analysis. However, these approaches assumed evenly and identically distributed data across nodes. Consequently, statistical procedures were derived to accommodate uneven node sample sizes and heterogeneous data distributions across nodes. Workflows and detailed algorithms were developed to highlight information-sharing requirements and operational complexity.

## 1 Introduction

### 1.1 Health Research at Scale

Learning health systems (LHS) are coming of age and are being deployed to address important health challenges at different scales. The framework starts by leveraging health data created across various activities. It obviously includes data points from clinics and hospitals, but the perimeter of data required to meaningfully and optimally address important problems is much wider and includes research cohorts, biobanks, quantified self data, environmental exposures and social service delivery.

While some questions might be addressed at the scale of an individual organisation, LHS focus on systems interactions and often require the analysis of processes and outcomes from various organisations. For example, to fully understand a cancer care trajectory, multiple data sources from multiple organisations will need to be examined to cover all relevant aspects (both within the traditional health system, but also in the community). This often implies at least regional organisations or bigger (provinces, states, countries), like in the context of the Health Data Research Network Canada (HDRN) or Health Data Research UK. Similarly, comparing various approaches is often a fruitful way to identify the best approaches and understand what works, why, and how to scale the promising projects. It can also be a way to amass a critical number of observations in the context of rarer diseases for example. Nevertheless, working with data from multiple data sources, from multiple organisations and located in multiple jurisdictions poses significant challenges.

Traditionally, the analytical methods used by researchers in the healthcare domain and others have relied on data pooling (sometimes referred to as data centralisation): all required data is physically copied to a single location where analysis can take place. However, when working with data from multiple jurisdictions (even when part of the same country like the Canadian provinces and territories), data pooling is often very difficult if not impossible for ethical, legal and social acceptability reasons.

There is, therefore, a pressing need to offer analytical methods allowing the analysis of such data without requiring the need to physically copy the data in a central location.

### 1.2 Distributed Analysis

More formally, this paper is concerned with frameworks where the data needed for a statistical analysis consists of the data about *n* individuals (referred to as the *analytical dataset*), which are not all stored in a single source but are partitioned among *K* locations which will be called *nodes* hereafter. The mereological sum of all the data held at each node therefore forms the analytical dataset. Data can be partitioned horizontally or vertically (or in a mixed way).

- A horizontal partition implies that all data pertaining to a given individual can be found in a single node. If we assume that patients receive care only in one province, Canadian provincial health administrative datasets hosted by organizations like Population Data BC, ICES in Ontario or the Manitoba Centre for Health Policy (MCHP) in Manitoba can be part of a horizontal partition. A clinical trial where each recruiting site captures all data for a given subject is another example.
- A vertical partition occurs when all data of a certain type is available in a single node for a group of individuals. A classic example is a hospital with its various information systems. All pathology results can be found in the pathology system, all billing information can be extracted from the finance system, all X-rays are accessible in the picture archiving and communication system (PACS), etc. But to get the full picture of the care received by a patient, multiple systems need to be interrogated. Similarly in the research setting, health administrative data may be in a provincial data centre and genomics data could be held in a research institute.

A mixed partition occurs when both principles partly apply: some individuals may have their data spread out across nodes, and different individuals may be present in different nodes.

#### 1.2.1 Assumptions

The difficulties in conducting analyses on a large scale mentioned above are often associated with horizontally partitioned data, and the current work focuses on this type of partition. The methods presented in this article might therefore not be directly applicable to vertically partitioned data.

One group of approaches often labelled as *distributed analysis* involves calculations at each participating node and exchanges of the resulting aggregated statistics with a *coordinating centre* (CC), which can itself also perform additional calculations based on the received aggregated statistics. The CC can be an organisation not responsible for a data node or a data node taking the additional role of CC for a given analysis.

It is important to note that whether in the more traditional way of data pooling or using distributed approaches (where the data is not copied centrally), data sources will be different on multiple levels. They will represent information using data models with significant variability in terms of structure and technology, but also in terms of semantics. This situation also leads to heterogeneous data where the presence of predictors and outcomes is likely to be different in different nodes. Different approaches (e.g. data mediation or extract-transform-load) have been developed to address these issues, and the current work assumes that one of them has been applied so that the data nodes mentioned hereafter are assumed to share the same structure, the same technological syntax and the same semantics as well as no missing data.

#### 1.2.2 Horizontally Partitioned Statistical Analytics

In what follows, the field that pertains to the statistical analysis of horizontally partitioned and semantically homogeneous data that cannot be consolidated into a central location will be called *Horizontally Partitioned Statistical Analytics* (HPSA).

Methodological contributions to this field have arisen from several streams of literature. Meta-analysis and meta-regression methods (see e.g. [45]) can be viewed as part of HPSA, e.g. by considering that each node-specific dataset belongs to a different “study”. However, their scope is narrower compared to HPSA because they typically assume that only established study-level estimates are available as data. Conversely, HPSA allows for the sharing of additional summary statistics between the nodes and the CC, such as gradients and Hessians to ensure the best possible performance at the global level. Since meta-analysis does not leverage any supplementary information that could be obtained from studies with access to patient-level data, it can be susceptible to biased estimation, especially in settings with rare outcomes or in the presence of data nodes with limited sample sizes [14]. As meta-analysis and meta-regression methods have been extensively covered in the literature, approaches specifically designed for the analysis of already-established study-level estimates will not be discussed hereafter.

An important research community that has generated a significant amount of analytical contributions is concerned with the massive data setting. There, a dataset often cannot be processed by a single server and is therefore split across multiple machines, which are then considered as nodes able to perform computations and send aggregated results to a CC tasked to fit a global model from them. The methodological avenues proposed in this setting share similarities with the ones designed for the multi-research facility setting involved in LHS, but also have important differences. For example, in the massive data setting, the experimenter has control over the distribution of individuals across nodes, which is typically not the case in multi-research facility studies. So while these approaches share mechanistic similarities and have been suggested as options to consider in the healthcare domain, some hypotheses may not hold. In regression settings, it is often reasonable to assume that the regression link between the response and covariate predictors is the same across nodes. However, assuming that the sampling distribution of covariates involved is equal across nodes is unrealistic in healthcare, particularly due to the presence of data centres that may systematically involve different types of patients. For example, certain clinics may predominantly serve older individuals. While this may not affect the estimation of parameter values, it can have implications for computing confidence intervals to ensure the validity of inferences.

So far, two reviews discussing methods applicable to horizontally partitioned data have been published in the literature [18][22]. However, their focus is on the massive data setting, which works almost invariably under the assumption of even sampling distribution of covariates and equal sample sizes across nodes, and statistical inference tasks beyond parameter estimation are barely covered. This makes them less helpful for healthcare research purposes since most studies involving data analyses rely on confidence intervals or hypothesis testing in settings where predictors’ distribution and sample sizes vary across nodes.

### 1.3 Contemporary challenges in HPSA

The problem is threefold. First, there is a need to raise awareness regarding the existence of HPSA approaches among researchers aiming at undertaking statistical analyses from horizontally partitioned data, especially in healthcare. The reflex is often to request data pooling because it is perceived as the sole option. This has been the tendency of requests made by researchers to to HDRN Canada. Practitioners are usually concerned with finding the most appropriate statistical model that will take into account as many of the features of their specific context of application as possible. Consequently, a clear and unifying mapping of the state of the HPSA field is needed for them to be informed of the scope of existing methods available for their analyses to see whether alternatives to pooling exist.

Second, as underlined above, methodological contributions came from research fields whose working assumptions can be fundamentally different from the ones researchers would be willing to assume in healthcare research. To ensure proper use of statistical inference techniques, it is necessary that the underlying assumptions of existing methods be adequately identified and understood. If necessary, these methods should be adapted to suit the specific requirements of healthcare applications, thereby ensuring accurate and reliable results.

Third, data custodians have to be properly informed on data-sharing requirements entailed by the use of a specific HPSA method applicable to a given research setting. While HPSA avoids the complexities of pooling data, there are still flows of information that have to be acceptable to data stewards. However, even in basic statistical scenarios, available methods are often presented in a way that makes them challenging to compare in terms of information-sharing requirements and operational complexity. Therefore, there is a need for clearer and more accessible presentations of these methods to facilitate decision-making regarding data sharing and operational implementation.

Although it would be ideal to offer managers a comprehensive operational workflow for each identified method to evaluate the information shared and execution complexity, with their accompanying underlying modelling assumptions, the abundance and diversity of available approaches make it unfeasible to accomplish this in a single paper. In fact, methods often differ in terms of their targeted application beyond their distributed aspect. For example, differences may exist in the studied model (e.g., linear, logistic or Cox regression, additive models), the dimensionality/sparsity of the predictor variable space, use of regularization or shrinkage, the presence of missingness, confounders, imbalances, heterogeneity, etc.

#### 1.3.1 Objectives

The objectives of this article are:

O1 To identify and map, from the literature, methodological approaches that make it possible to perform confidence intervals estimation and hypothesis testings from a horizontally partitioned dataset;

O2 Among the approaches identified, to describe the ones that allow to conduct general linear model analyses, and to identify their distributional assumptions;

O3 Based on the approaches identified for GLM-based inferences, to present methods adapted to the setting of uneven sampling distributions across nodes, and to compare them in terms of information-sharing requirements and operational complexity.

A scoping review methodology was chosen to achieve objective O1 of mapping the state of the field of HPSA that pertains to inference procedures. For our second objective (O2), we identified, from the articles selected from the literature search, the ones that presented a methodological framework for conducting statistical inference procedures from a GLM with horizontally partitioned data. We then used these frameworks to derive and describe GLM estimators that are applicable to horizontally partitioned datasets. For each identified method, we analyzed and reported its communication workflow and the distributional assumptions. For our third objective (O3), we first used statistical theory to adapt the identified procedures to the unequal sample size and uneven covariate distribution setting. Algorithms and mathematical expressions for the quantities involved are reported. For conciseness, we present mathematical formulas for estimation procedures of confidence intervals only. Expressions involved for hypothesis testings are similar and can be deduced following the close connecting between confidence intervals and hypothesis tests in GLMs, see e.g. [1].

The mathematical description of the GLM setting considered for this analysis is described below, along with mathematical notations to be used.

### 1.4 Mathematical framework

In the following, lowercase bold letters will represent vector-valued quantities, while uppercase bold letters will denote matrices. The *j*^th^ element of any vector ***a*** *∈* ℝ^*p*^ will be denoted as [***a***]_*j*_. Similarly, the entry at position (*j, l*) of any matrix ***A****∈* ℝ^*p×p*^ will be denoted as [***A***]_*jl*_. If *g* is a real-valued and invertible function, we will use *g*^*−*1^ to represent its inverse. Additionally, if *f*_***θ***_ is a real-valued function that depends on a parameter vector ***θ*** and is twice continuously differentiable, *∇*_***θ***_ *f*_***θ***_ and 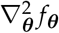 will respectively indicate the gradient and Hessian matrix of *f*_***θ***_ with respect to ***θ***.

#### 1.4.1 Model mathematical assumptions

A mathematical depiction of the horizontally partitioned data framework studied in this paper is as follows. There are *n* individuals horizontally partitioned across *K* data storage nodes. Each node’s dataset is denoted by 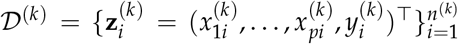, where 1 *≤ k ≤ K*. Here, 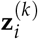 represents the measurements on the *i*^th^ individual at node *k*, where 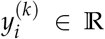 denotes their response variable and 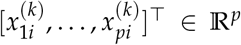 denotes their covariate vector. The total sample size at node *k* is denoted by *n*^(*k*)^. The combined datasets 𝒟 ^(1)^, …, 𝒟 ^(*K*)^ make up the whole dataset without any duplicated individuals, indicating that 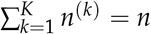.

Throughout the analysis, it is assumed that the 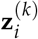 ’s are independent across 1 *≤ i ≤ n*^(*k*)^ and 1 *≤ k ≤ K*, and there is no missing data. Additionally, the size of the covariate space (i.e., the dimension of 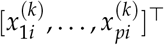, which is equal to *p* representing the number of features to include as predictors in the GLM) is assumed to be low, eliminating the need for regularization or variable selection. Finally, it is assumed that each node possesses a non-negligible proportion of the whole dataset. Specifically, for each *k ∈* {1, …, *K*}, the quantity *n*^(*k*)^/*n* is bounded away from 0 and 1 as the sample size *n* tends to infinity, denoted as *n*^(*k*)^/*n → p*^(*k*)^ *∈* (0, 1).

#### 1.4.2 Mathematical description of the GLM framework

The formulation of the GLM considered in this article encompasses various commonly used regression models such as linear regression, logistic regression, Poisson regression, and probit models. It assumes that the density or probability mass function of each response variable (known as the random components) belongs to the exponential family of distributions. Within this formulation, the (conditional) mean of the response variable is expressed as a function of a linear combination of the corresponding covariate vector. Formally, it assumes that there exist unknown parameters ***β***^*⋆*^ *∈* ℝ^*p*+1^ and *ϕ*^*⋆*^ *>* 0, and known model-specific functions *b, c, g, h* such that with 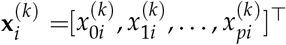 and 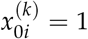,

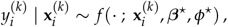

where, for any ***β*** = [*β*_0_, *β*_1_, …, *β*_*p*_]^*⊤*^ *∈* ℝ^*p*+1^ and *ϕ*,

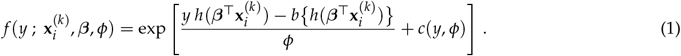

In formula (1), *b* maps real numbers to real numbers and is such that 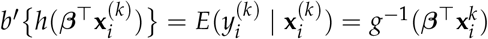, with 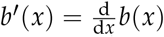. In this framework, *g* is called *link function*, the term 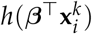 is usually referred to as the *natural parameter* and *b* as the *cumulant* function. *ϕ* is often called the *dispersion parameter*, and is either known (e.g. with *ϕ* = 1) or unknown. When *h*(*x*) = *x* (i.e. *h* is the identity function), the link *g* is called *canonical*.

The logistic regression model is obtained upon taking *ϕ* = 1, *h*(*x*) = *x, b*(*x*) = log(1 + *e*^*x*^), *c*(*y, ϕ*) = 0 and *g*(*x*) = log {*x*/(1 *− x*)}. The linear regression model with homoskedastic residual error variance *ϕ* is derived upon setting *h*(*x*) = *x, b*(*x*) = *x*^2^/2, *c*(*y, ϕ*) = *− y*^2^/(2*ϕ*) − log(2*πϕ*)/2 and *g*(*x*) = *x*. Hence, both the logistic and the linear regression models rely on a canonical link function in the exponential family distribution.

## 2 Materials and Methods

### 2.1 Methodology related to objective O1

Scoping reviews are well-suited to efficiently map key concepts within a research area [2]. They are widely acknowledged for their ability to clarify working definitions and conceptual boundaries in a specific topic or field [39], facilitating a shared understanding among researchers regarding the status of the research area. These considerations make the scoping review methodology well-designed to achieve objective O1.

Scoping studies utilize systematic searches of relevant databases, employing specific keywords to define the boundaries of the research field. However, identifying these keywords can be challenging, particularly when relevant papers are scattered across different research streams or in independent clusters that do not reference each other. To address the risk of overlooking significant methodological contributions due to a limited number of keywords, a snowballing literature search was initially conducted to generate a comprehensive list of keywords related to HPSA. The scoping review then proceeded with a systematic literature search using the identified keywords. It’s worth noting that, since the planning of the scoping study is independent of the search approach, the guidelines presented in [2] are still appropriate.

#### 2.1.1 Methodology pertaining to the snowballing keywords search

Snowballing is generally used as a literature search method aiming at identifying papers belonging to a given field [54]. It typically consists in three steps:

1. Initiate searches in prominent journals and/or conference proceedings to gather an initial set of papers.
2. Conduct a backward review by examining the reference lists of the relevant articles discovered in steps 1 and 2 (continue iterating until no new papers are found).
3. Perform a forward search by identifying articles that cite the papers identified in the previous steps.

To avoid selection bias, the initial set of papers for the snowballing approach in (1) is sometimes generated through a search in Google Scholar (see e.g. [23]). The latter strategy was used here too.

As mentioned earlier, here, the snowballing search strategy was used in preparation for the application of the scoping review protocol, with the goal of identifying relevant keywords. Specifically, the starting set of papers was assembled by screening titles and abstracts from the first 50 papers generated through a Google Scholar search using the strings *distributed inference* and *federated inference*. The main inclusion criterion was “presents, applies or discusses a statistical inference method to analyse horizontally partitioned data”. Then, the backward and forward snowballing steps approaches were applied.

From the set of keywords found in the selected papers, a list of those relevant to HPSA but not directly associated with any specific method was retained for the scoping review step. It is worth noting that, since the objective of the scoping review is to identify statistical inference methods for horizontally partitioned data, keywords linked to method identifiers have to be excluded from the retained list to avoid pre-selection bias in the scoping review phase of this project.

Selected keywords that were identified from the snowballing literature search are *distributed algorithms, distributed estimation, distributed inference, distributed learning, distributed regression, federated inference, federated estimation, federated learning, privacy-protecting algorithm, privacy-preserving algorithm* and *aggregated inference*.

#### 2.1.2 Methodology pertaining to the scoping review

The scoping review’s methodological framework of Levac et al. [26] (see also [2]) was followed. The steps are briefly described below. A detailed protocol is available in Appendix A.

##### Search strategy

We conducted a comprehensive search across four bibliographic databases, namely (1) MEDLINE, (2) Scopus, (3) MathSciNet, and (4) zbMATH, to encompass the interdisciplinary nature of the topic and identify relevant research articles. Our research strategies were based on two key concepts: distributed data and statistical inference. In addition to the keywords obtained from the snowballing step, we incorporated terms like *confidence interval* to target articles focusing specifically on statistical inference. To ensure the inclusion of recent advancements, our search was limited to papers published from 2000 onwards. This cutoff date was chosen to account for the emergence of distributed data, the prevalence of massive datasets, and advancements in technology. It was set conservatively to capture any early-developed methods and ensure comprehensive coverage of the topic.

##### Selection process

After completing the primary research, a two-stage selection process was employed. Initially, two authors (MPD, FCL) collaborated to screen all articles identified through the research strategy based on their titles and abstracts. Subsequently, the full texts of the selected articles were independently reviewed by both authors to finalize the selection process. This rigorous approach ensured a thorough evaluation of each article’s relevance and eligibility for inclusion.

The primary inclusion criterion for the selection process was as follows: *Presents a solution for conducting inferential statistics on horizontally partitioned data*. This criterion was utilized to ensure that the chosen articles specifically addressed the methods associated with performing statistical inference on horizontally partitioned data.

The following exclusion criteria were derived directly from objective O1:

- Does not address inferential statistics, including confidence intervals, hypothesis testing, or asymptotic normality.
- Does not provide a methodological contribution.
- Presents a solution for encryption or secret-sharing.

To ensure the inclusion of validated approaches, the selection process only considered published papers that had full-text availability in English or French. Discussion papers were excluded as they do not present novel methods or approaches.

Exclusion was considered if any of the exclusion criteria were met or if any of the inclusion criteria were not met. Finally, the references of each included article from the databases were assessed to identify any relevant articles that may not have been captured during the initial screening due to specific keywords. This additional step in the selection process was necessary given the broad range of vocabulary used to describe applicable approaches in our context.

##### Data extraction and analysis plan

Data extraction for the included articles was conducted by one author (MPD) and followed a collectively developed data-charting form. Model type (*parametric regression, semi-parametric regression, non-parametric regression* or *not specific to regression*) and number of communication from CC to nodes (0 or *≥* 1) were among the data extracted. All methods from the included articles were subsequently classified according to their specified characteristics, as outlined in the protocol. Additionally, as part of the analysis, we conducted a screening of the general distributed approaches commonly employed across all specific methods.

### 2.2 Methodology related to objective O2

To achieve objective O2, three steps were taken. First, we identified methodological approaches from articles included in the scoping review that enable parameter and confidence interval estimations from horizontally partitioned data within a standard GLM framework. Methods designed specifically for the particular cases of linear or logistic regression were also reported but were not analyzed in detail. Second, we extracted workflows for each approach to determine the information exchanged between data storage nodes and the CC. Third, we analyzed the mathematical assumptions necessary for parameter estimation and the consistency of confidence interval procedures. We specifically reported the assumptions related to the distribution of node-specific covariates.

#### 2.2.1 Identification of the approaches

To identify approaches that enabled the fitting of any GLM using horizontally partitioned data, two authors (FCL, MPD) independently assessed all articles included in the scoping study. The reviewers specifically looked for articles that discussed approaches applicable to the GLM class described in section 1.4, including likelihood-based methods, M-estimation, and estimating equations. Additionally, we identified and reported articles that specifically focused on regression settings for linear or logistic regression. However, unless the method described was considered easily adaptable to the GLM framework, these articles were not retained for detailed analysis.

A method was selected if it provided an algorithm for fitting GLMs using horizontally partitioned data, aligning with the characteristics outlined in section 1.4. In cases where an article presented asymptotic normality results for the estimators but did not provide an estimator for the asymptotic variance-covariance matrix, the article was still retained, and an estimator for the asymptotic variance was derived using the available calculated quantities.

Since the GLM framework in section 1.4 assumes no missing values, low dimensionality, and a small number of nodes relative to the total sample size, any terms related to these specific conditions mentioned in an article’s methodology were disregarded. Consequently, the calculations for confidence intervals were adjusted accordingly. If an article solely focused on one of these aspects without contributing to the overall methodology, it was not included in the final selection.

Methodological components regarding parameter estimation and confidence interval procedures were extracted from the screened articles. Specifically, the focus was on understanding how parameters should be estimated within a horizontally partitioned framework and how confidence intervals should be computed for these parameters. For each article, the formulas related to quantities shared among the nodes and quantities calculated by the CC were derived and analyzed. These formulas were examined within a workflow that indicated the necessary circulation of information for the procedure to be executed.

##### Reported results

The rationale behind each method that was deemed suitable for fitting GLMs was documented, along with the corresponding reference to the paper included in the scoping study where the method was introduced or discussed.

Articles that discussed approaches specifically applicable to the cases of linear or logistic regression were also mentioned, but not elaborated on in detail.

### 2.3 Methodology related to objective O3

In most statistical settings with horizontally partitioned data, it is commonly assumed that the sample sizes of the data nodes are equal and that the distribution of covariates is the same across all nodes. However, when the number of nodes is fixed and relatively small compared to the sample sizes, it is possible to adapt a particular approach to handle situations where the sample sizes and covariate distributions vary across nodes. This can be achieved by combining the theoretical arguments presented in the original article of the method with the principles of asymptotic statistics theory concerning maximum likelihood estimation.

To adapt a given approach for situations where sample sizes and covariate distributions differ across nodes, the following steps were taken:

1. The formulas for the relevant quantities were modified to emphasize the changes caused by this scenario. It was ensured that the adapted quantities were equivalent to their counterparts presented in the original article for an equal sample size setting.
2. Using asymptotic theory, an asymptotic normality result was derived for the estimators of interest, assuming a set of assumptions that accommodated potential variations in sample sizes and covariates sampling distribution across nodes, while still enabling meaningful theoretical arguments.
3. Formulas for the asymptotic variances were derived. Statistical theory on maximum likelihood estimation was employed to obtain consistent estimators for asymptotic variances. The latter estimators were derived under the constraint that they had to be calculated without requiring any additional communication round between the CC and the nodes. Thus, throughout the adaptation process, the communication workflow remained unchanged compared to the original method.

These steps ensured the mathematical correctness of adapting the approaches to handle different sample sizes and covariate distributions across nodes. Importantly, the adaptation maintained consistency with the original method’s communication workflows.

#### Statistical estimates of interest

A standard GLM typically includes one or two unknown parametric components. The first are the ***β*** parameters, which are commonly assumed to be unknown. The second parameter is the nuisance parameter *ϕ*, which can either be known (e.g., in logistic models) or unknown (e.g., in linear models). In practical applications, when *ϕ* is unknown, its estimated value is often not the main focus, although the latter is necessary to estimate the asymptotic variance of the ***β*** parameter estimates.

In the upcoming analysis, we will assume that the parameter *ϕ* is unknown and estimated using the recommended approach in the selected methods. However, in the case where *ϕ* is known, the process becomes simpler. This involves substituting the known value of *ϕ* and disregarding the estimation step. It is important to highlight that estimating *ϕ* requires additional information to be shared between the nodes and the CC, but it does not necessitate any extra communication round between them.

The estimation process for both the ***β*** parameters and the *ϕ* parameter are discussed. Additionally, we explained how to compute an estimator for the asymptotic variance specifically for the estimator of ***β***^*⋆*^. It is important to note that the results presented below can be modified and extended to develop a similar procedure for estimating *ϕ*^*⋆*^.

Using these results, based on an estimator of ***β***^*⋆*^, say 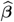 and a formula for the estimator of the asymptotic variance-covariance matrix involved in its associated asymptotic normality result, say 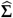, Wald-type (1 *− α*) confidence intervals can be computed for each component of ***β***^*⋆*^ using the formula

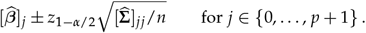

#### Reported results

For each approach considered, we presented the formulas necessary to compute the final estimates of the ***β*** parameters and their corresponding confidence intervals. The presentation of these formulas was designed to emphasize the communication workflow. Furthermore, a comprehensive algorithm was provided, outlining the step-by-step process.

In addition, the asymptotic normality of the ***β*** parameter estimators was stated, accompanied by the formula for the asymptotic variance and its consistent estimator. Detailed proofs for these results can be found in the Appendix B.

## 3 Results

### 3.1 Results related to objective O1

#### 3.1.1 Search outcomes from the scoping review

As presented in Figure 1, a total of 1407 articles were initially identified across all four databases after removing duplicates. Subsequently, a majority of these articles (n=1274) were excluded based on the evaluation of titles and abstracts, leaving 133 articles for eligibility assessment through full-text review. Following this assessment, 29 articles were included from the databases. Additionally, by reviewing the references of the included articles, 12 more articles were identified and added to the study.

**Figure 1.**
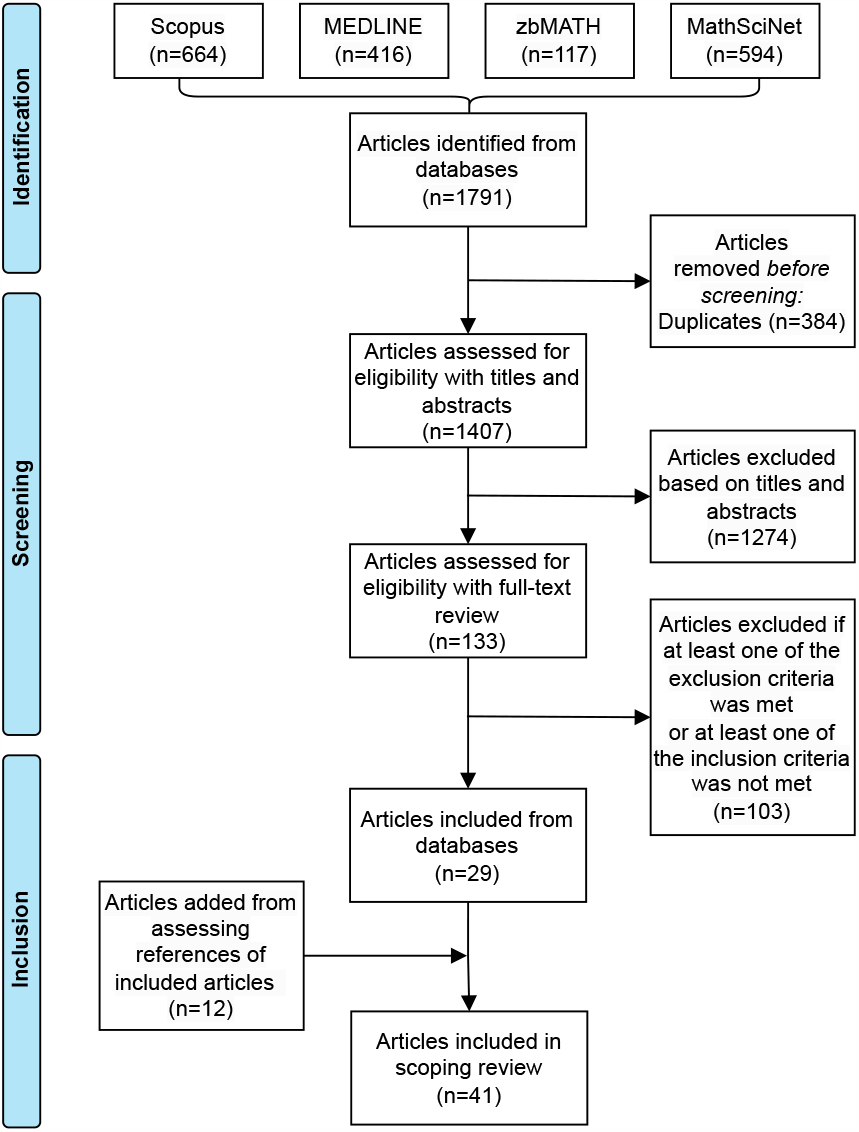
Article selection process for the scoping review. Detailed inclusion and exclusion criteria are described in the text and in the protocol.

Among the additional 12 articles obtained through the assessment of references from included articles, it is observed that most of them did not mention statistical inference or related terms in their abstracts (e.g., [14][15][41]). Consequently, these articles were not captured in the initial database search results. Furthermore, some articles directly referred to the specific method used without including any keywords related to horizontally partitioned data in their abstracts or titles (e.g., [4][11]), which greatly reduced the chance of initially identifying them. However, during the process of reviewing the references of included articles, all the relevant papers that were initially identified through the snowballing strategy were eventually retrieved either through the search strategy or the selection process based on the references of included articles.

#### 3.1.2 Results of the scoping review

Each article included in the scoping review put forth one or multiple methodological approaches pertaining to objective O1. Similarities are differences regarding the communication schemes involved and their background of origin are summarized below.

First, all selected articles discuss one or more statistical procedures that operate on horizontally partitioned data using one of the communication schemes depicted in Figures 2 to 5.

**Figure 2.**
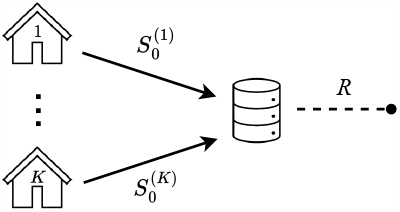
Workflow I.

- In Workflow I, as shown in Figure 2, each node calculates summary statistics from its own samples, and the results are sent to the CC. The CC combines the information provided by each node to produce the final estimates. This communication approach is commonly referred to as a “one-shot” or “non-iterative” in the literature, although not always consistently.
- In Workflow II, as shown in Figure 3, multiple communication rounds are allowed between the CC and the data storage nodes. This allows for iterative interactions between the nodes and the CC to refine the estimates.

**Figure 3.**
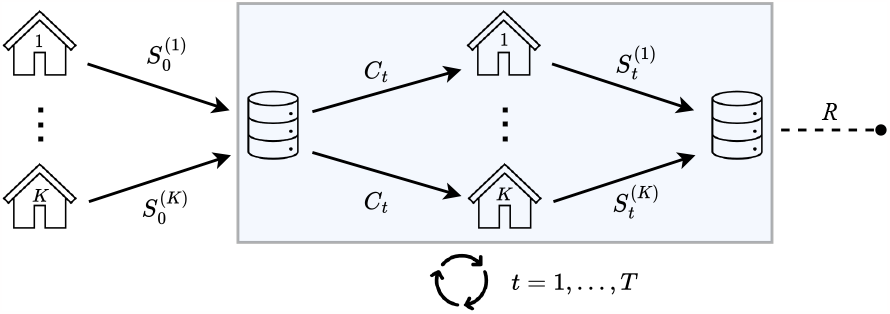
Workflow II.
- Some approaches fundamentally differ from the two previous workflows by assigning a different role to one of the nodes, say node 1, compared to the others. These approaches operate using Workflow III as illustrated in Figure 4, where node 1 follows a distinct communication pattern compared to the other nodes. In the papers included in the scoping review that discuss these approaches, node 1 is invariably designated as the CC. However, in the context of the current paper, their roles were distinguished. The additional step performed by the CC, which involves data aggregation, can be particularly well-suited for privacy protection purposes in practice.

**Figure 4.**
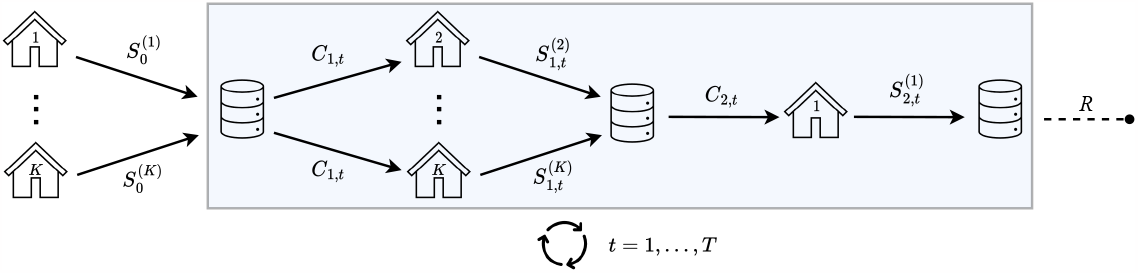
Workflow III.
- The particular setting shown in Workflow IV in Figure 5 requires two back-and-forth communication exchanges between each node and the CC at each iteration. This communication pattern distinguishes it from the other workflows.

**Figure 5.**
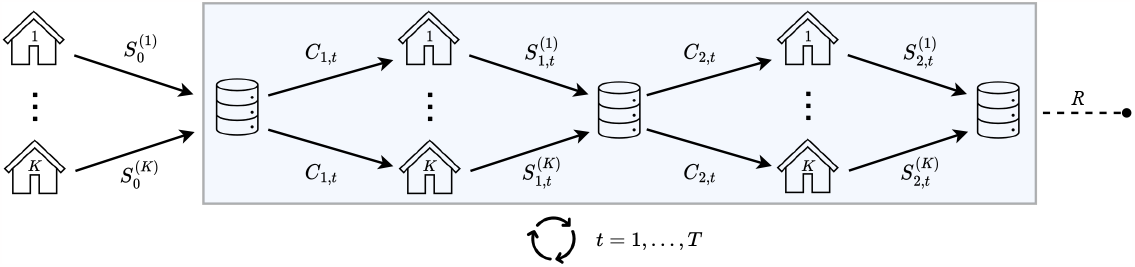
Workflow IV.

In light of the preceding discussion, from an operational standpoint, two categories of workflows emerge. On one hand, there are workflows that do not necessitate any communication from the CC to the nodes, which are captured in Workflow I. On the other hand, there are workflows that involve one or more communication exchanges from the CC to the nodes, which are captured in Workflows II, III and IV.

In order to emphasize similarities among the methods presented in these articles and facilitate the identification of methods suitable for specific purposes, a systematic classification is presented in Table 1. The articles are categorized based on the type of models employed and the number of communications from the CC to the individual nodes.

**Table 1:** Classification of articles included in the scoping review.

| Type of model | 0 communication from CC to nodes | $\geq 1$ communication from CC to nodes |
| --- | --- | --- |
| Parametric regression | Basiri, Ollila and Koivunen [5]; Battay et al. [6]; Fan, Guo and Wang [17]; Guo, Sun and Jiang [19]; Chen and Xie [11]; Lin and Xi [29]; Rosenblatt and Nadler [41]; Zhang, Duchi and Wainwright [60]; Chang, Bu and Long [9]; Wu et al. [56]; Hector and Song [20] | Huang and Huo [21]; Jordan, Lee and Yang [24]; Mozafari-Majd and Koivunen [35][36]; Yue, Kontar and Gómez [58]; Duan, Ning and Chen [13]; Duan et al. [14]; Tong et al. [47]; Di, Wang and Lian [12]; Edmondson et al. [16]; Luo and Li [33]; Shu, Young and Toh [44] |
| Semi-parametric regression | Zhao, Cheng and Liu [61]; Park et al. [38] | Luo, Sun and Zhou [32]; Duan et al. [15] |
| Non-parametric regression | Liu, Shang et Cheng [30]; Zhang et al. [59]; Volgushev, Chao and Cheng [51] | Wang et al. [52] |
| Not specific to regression | Atta-Asiamah and Yuan [3]; Minsker [34]; Lin and Xi [28]; Bruce et al. [8]; Chen and Peng [10]; Nezakati and Pircalabelu [37]; Banerjee, Durot and Sen [4]; Shi, Lu and Song [43]; Wu et al. [55] | Lai, Hanning and Lee [25] |

The majority of the methods were published within the methodological setting of Big or Massive data/Multi-machine, while some were reported within the context of healthcare research. Within the Big or Massive data/Multi-machine methodological setting, many methods involve an initial step of random data partitioning among multiple machines. However, certain methods assume a scenario where data is already stored on separate machines, as observed in [17] and [24]. Furthermore, it is worth noting that no articles published prior to 2010 were included, aligning with our initial hypothesis regarding the identification of contemporary methodological settings. The majority of the included articles (30 out of 41) were published after the year 2018.

The majority of articles address a setting where a CC exists external to the nodes, as exemplified by articles such as [28], [51], and [58]. In contrast, as mentioned above, some articles designate one of the nodes to assume this central role, as demonstrated in [9].

The methods identified through our search strategy share a common characteristic of utilizing a global model that incorporates population-level parameters. In some cases, these parameters may also include node-specific components to accommodate node-specific statistical heterogeneity in the outcome-predictors relationship, which captures deviations from the population-level conditional probability distribution of the outcome given the predictors.

A few of the reported methods have the capability to yield results identical to those obtained if the individual line data were pooled from all nodes, see e.g., [56] and [44].

### 3.2 Results related to objective O2

Six approaches were selected as applicable to the standard GLM framework discussed in 1.4. They all assumed that nodes had equal sample sizes and identical distributions for the covariates.

#### 3.2.1 Simple averaging

One of the simplest methods for horizontally partitioned data analysis, often referred to as the “simple averaging method” or the “divide-and-conquer” approach, has been extensively studied in the literature, see [60] and [41] which were included in our scoping study. It operates through Workflow I in Figure 2. In this approach, node-level model estimates are gathered and averaged at the CC to generate the final estimates.

In the context of GLM, each node is initially tasked with calculating the maximum likelihood estimator (MLE) of the ***β***^*⋆*^ and *ϕ*^*⋆*^ parameters using their respective data. Additionally, the Hessian matrix of the log-likelihood function with respect to the ***β*** parameters must be computed for constructing Wald-type confidence intervals. The estimated parameters and the computed Hessian matrix are then transmitted to the CC.

The final parameter estimates of ***β***^*⋆*^ are obtained by averaging the node-specific estimates, while the local Hessians and estimates of *ϕ*^*⋆*^ are utilized to compute an estimator for the asymptotic variance.

#### 3.2.2 Single distributed Newton-Raphson updating

The single distributed Newton-Raphson updating method is an iterative procedure that includes an additional communication round between the CC and the nodes, compared to the simple averaging method. It was originally proposed as the “distributed one-step” method in [21], but here it is referred to by a different term to avoid any confusion regarding communication complexity. This method operates using Workflow II, as depicted in Figure 3, with *T* = 1 (where *T* represents the number of cycles in the iteration scheme). It enhances the simple averaging estimators by incorporating a single distributed Newton-Raphson updating step.

In the context of GLM, each node first calculates the MLE of ***β***^*⋆*^ and *ϕ*^*⋆*^, and transmits them to the CC. The CC aggregates these estimates using averaging and sends the result back to the nodes. The nodes then compute the gradient and the Hessian matrix of the log-likelihood function, evaluated at the received ***β***^*⋆*^ and *ϕ*^*⋆*^ estimates. Subsequently, the gradient and the Hessian matrix are sent back to the CC, which averages them and computes a Newton-Raphson updating step based on the simple averaging estimates. An estimator for the asymptotic variance can be calculated by utilizing the received Hessian matrices and the updated estimate of *ϕ*^*⋆*^.

#### 3.2.3 Multiple distributed Newton-Raphson updatings

The multiple distributed Newton-Raphson updating method leverages the fact that, for standard GLMs, the algorithm typically used to calculate the MLE of ***β***^*⋆*^ and *ϕ*^*⋆*^ in a centralized pooled setting can be executed in a distributed manner without any loss of information. This is possible because the algorithm relies on Newton-Raphson updatings (or sometimes Fisher scoring updatings) that are expressed using two sums of node-specific summary statistics, namely local gradients and local Hessian matrices of the log-likelihood function, evaluated at the ***β***^*⋆*^ and *ϕ*^*⋆*^ estimates from the previous iteration. A version of this method is proposed in [56] under the logistic regression framework. It operates through Workflow II in Figure 3 for a general *T ≥* 1.

#### 3.2.4 Distributed estimating equation

The class of estimating equations estimators is vast and encompasses a broad range of statistical estimation techniques, including likelihood-based approaches that rely on searching for critical points. The fundamental idea behind estimating equations methods is to establish a system of equations that involve both the sample data and the unknown model parameters. These equations are then solved to determine the parameter estimates. MLEs, which are obtained by setting the gradient of the log-likelihood function with respect to the unknown parameters equal to zero, belong to the class of estimating equations estimators.

The distributed estimating equations approach involves gathering summary statistics from nodes at the CC level, enabling the reconstruction of the estimating equations, or more commonly, an approximation of them that would have been obtained in a pooled centralized setting. This method is discussed in [29] and operates using Workflow I, as depicted in Figure 2.

In the context of GLMs, the distributed estimating equations approach involves initially assigning each node the task of computing and sending their local MLEs and the Hessian matrix of their local log-likelihood, evaluated at those MLEs, to the CC. The CC utilizes these received quantities to reconstruct the global estimating equations or an approximation thereof. This reconstruction ultimately leads to an analytical solution for obtaining the resulting estimates. Confidence intervals are computed using a combination of the Hessian matrices and the final estimator of *ϕ*^*⋆*^.

It is important to note that when this approach is applied in the context of linear regression, it enables the acquisition of ***β***^*⋆*^ parameter estimates that are identical to those obtained in a pooled centralized setting.

#### 3.2.5 Distributed estimation using a single gradient-enhanced log-likelihood

This method differs fundamentally from the ones discussed thus far, as it involves a distinct role for one particular node in obtaining model parameter estimates. It operates using Workflow III, as depicted in Figure 4, and was proposed in [24] under the name “Surrogate likelihood”. This approach relies on an approximation of the global likelihood by viewing it as an analytic function. It expands the global likelihood into an infinite series around an initial guess 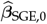 and replaces the higher-order derivatives (order *≥* 2) of the global likelihood with those of a Taylor expansion of a node’s (e.g., node *k* = 1) local likelihood around the same value. By following this procedure, the so-called surrogate likelihood can be solved using data from node *k* = 1 and aggregated gradients from nodes *k ∈* {2, …, *K*}.

In the context of GLM, the CC first collects the necessary information to compute initial estimates for the parameters ***β***^*⋆*^ and *ϕ*^*⋆*^. These initial estimates can be obtained through various approaches, such as a simple averaging estimator or the MLEs computed using data from node 1. These initial estimates are then transmitted to nodes *k∈* {2, …, *K*}. Each of these nodes calculates the gradient of the log-likelihood function, evaluated at the received estimates, and sends it back to the CC. The CC averages these gradients and sends the result to node Node 1 solves a gradient-enhanced log-likelihood using its own data and the received average gradient. The resulting estimate is sent back to the CC as the final estimate. To compute confidence intervals, each node must send the Hessian matrix of its local log-likelihood function, evaluated at the initial received estimate.

The steps related to estimation can be repeated multiple times.

#### 3.2.6 Distributed estimation using multiple gradient-enhanced log-likelihoods

This method is in the spirit of the *distributed estimation using a single gradient-enhanced log-likelihood* approach described above, except that all nodes have to solve a gradient-enhanced log-likelihood instead of only one of them. Results pertaining to statistical inference are discussed in [17] under a penalized setting. A non-penalized version of this method was introduced in [42] although the latter did not discuss confidence intervals or hypothesis testing, and hence was not included in our scoping review. It operates through Workflow IV depicted in Figure 5.

### 3.3 Results related to objective O3

In what follows, let the log-likelihood of the data stored in node *k* (using 𝒟 ^(*k*)^) be denoted by

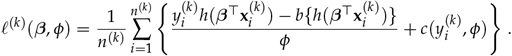

Also, let ***D***^(*k*)^(***β***) *∈* ℝ^*p*+1^ be such that

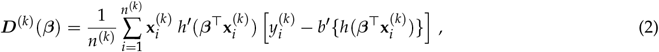

and define the (*p* + 1) *×* (*p* + 1) matrix ***V*** ^(*k*)^(***β***) as

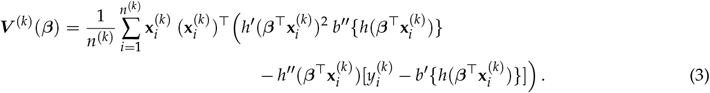

Since 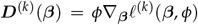, then, solving the equation ***D***^(*k*)^(***β***) = 0 yields the node-specific MLE of ***β***, denoted hereafter by 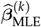. The matrix ***V***^(*k*)^ (***β***) is equal to *−∇*_***β***_ ***D***^(*k*)^ (***β***) and relates to Fisher information matrix through the equation 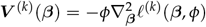.

Finally, set

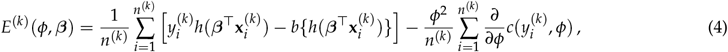

and

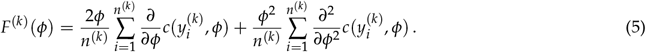

Because *E*^(*k*)^(*ϕ*, ***β***) = *−ϕ*^2^(*∂*/*∂ϕ*)*ℓ*^(*k*)^(***β***, *ϕ*), when *ϕ* is unknown, solving the equation 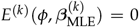 for *ϕ* yields its node-specific MLE of *ϕ*^*⋆*^. We have *F*^(*k*)^(*ϕ*) = *− ∂*/*∂ϕ E*^(*k*)^(*ϕ*, ***β***).

#### 3.3.1 Simple averaging

The simple averaging method follows upon execution of Algorithm 1. First, each data node computes their local maximum by solving successively ***D*** ^(*k*)^ (***β***) = 0 and 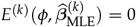. To compute the confidence intervals at the CC level, the entries of the (*p* + 1) *×* (*p* + 1) matrix 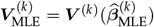. have to be computed from Formula (3) with 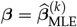 Then, the set

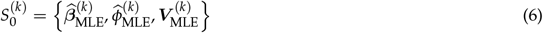

is sent to the CC. The parameter estimates are then aggregated by the CC through averaging. Specifically, the CC computes

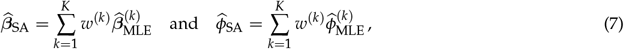

where *w*^(1)^, …, *w*^(*K*)^ are weights (i.e., *w*^(*k*)^ *≥* 0 and 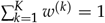) used to combine each node’s contribution. Often, weights can be taken proportional to local sample sizes, leading to the choice *w*^(*k*)^ = *n*^(*k*)^/*n*.

Wald-type confidence intervals for ***β***\* can be constructed based on the fact that the sequence 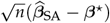 converges in distribution to a centred normal random variable with covariance matrix

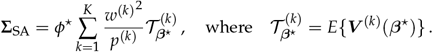

See Appendix B.3.3. Since 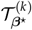 is consistently estimated by 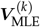 and *ϕ*^*⋆*^ by 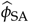, and as *p*^(*k*)^ can be estimated by *n*^(*k*)^/*n*, it follows that a consistent estimator for **Σ**_SA_ is given by

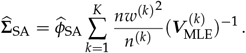

The simple averaging final estimates are then given by

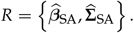

##### Algorithm 1

Simple averaging inference procedure

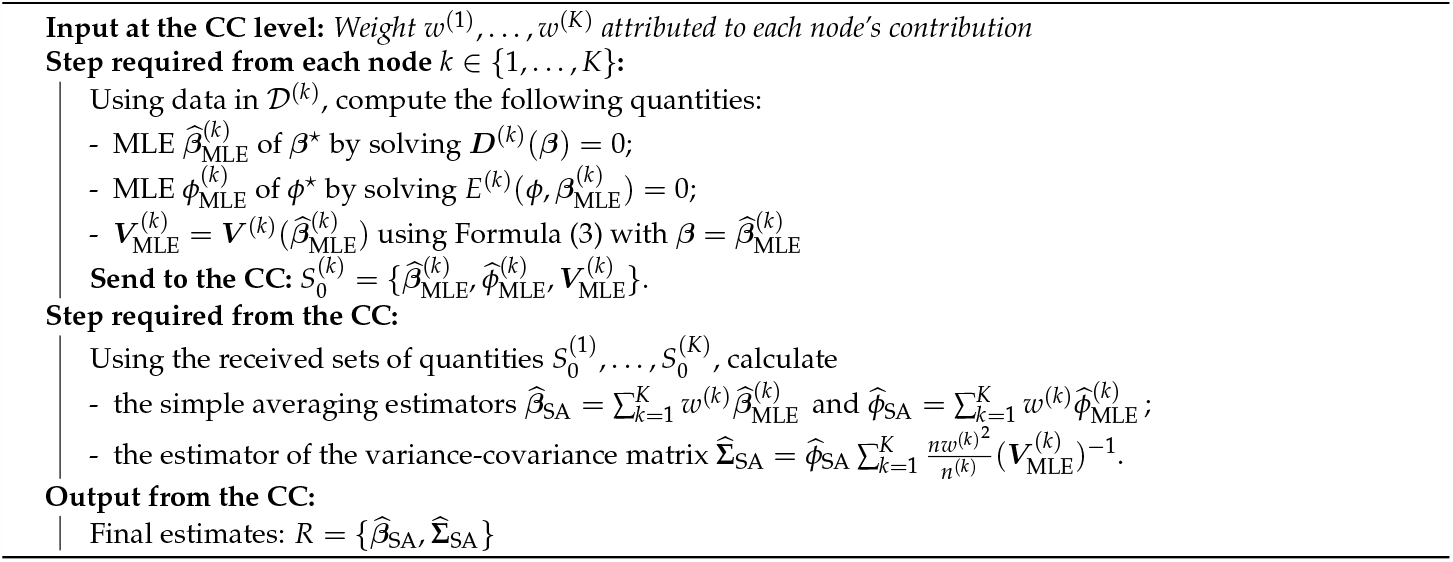

#### 3.3.2 Single distributed Newton-Raphson updating

The single distributed Newton-Raphson updating method follows upon execution of Algorithm 2 with *T* = 1. First, the CC gathers summary statistics to compute the simple averaging estimators of ***β***^*⋆*^ and *ϕ*^*⋆*^ without their accompanying confidence interval. Hence, for *k∈* {1, …, *K*}, and with 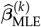 as above (6), node *k* sends to the CC the quantities

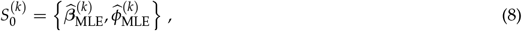

which uses them to compute 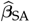 and 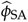 using the formulas in (7).

For reasons of convenience that will become clear later, the notation 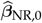 and 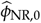 will be utilized instead of 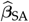 and 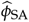, respectively. In this notation, the set of values

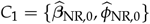

is broadcasted to data nodes, which are then tasked to compute and send back the quantities

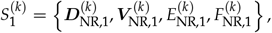

where for any integer *t ≥* 1, one defines

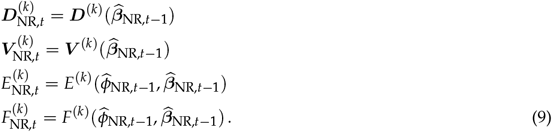

Upon receiving the 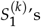 from each node, the CC calculates the following weighted averages:

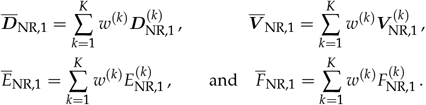

This enables the CC to execute Newton-Raphson updates from 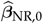 and 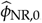, respectively:

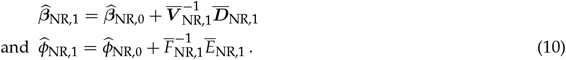

It is shown in Appendix B.3.4 that

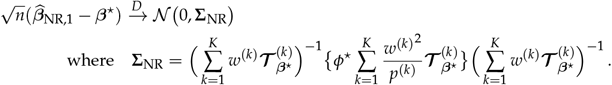

Since 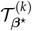 is consistently estimated by 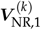 and *ϕ*\* by 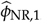, and as *p*^(*k*)^ can be estimated by *n*^(*k*)^ /*n*, it follows that a consistent estimator for **Σ**_NR_ is given by

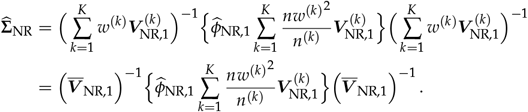

The method’s final estimates are then given by

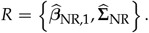

#### 3.3.3 Multiple distributed Newton-Raphson updatings

The multiple distributed Newton-Raphson updatings method follows upon execution of Algorithm 2 with *T >* 1. The first communication cycle follows the same procedure as described above for the single distributed Newton-Raphson updating method. It involves distributively computing a simple-averaging estimator and then performing a Newton-Raphson iteration starting from this estimator. The Newton descent is calculated as described in Equation (10).

Formally, the algorithm begins with each data node *k* sending the set of quantities 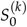 as described in Equation (8) to the CC. Next, the CC calculates the simple averaging estimators using Formula (7), and uses them to initialize 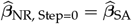 and 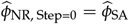.

The following steps are then repeated for a certain number of iterations. At iteration *t*, starting from *t* = 1, the CC broadcasts the values 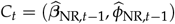 to the data nodes. The data nodes compute the quantities 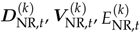 and 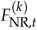 as defined in Equation (9), and send them back to the CC.

The CC then utilizes these quantities to perform a Newton update. Specifically, it calculates 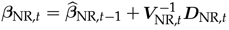 and 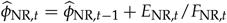.

If the iterative cycle is repeated until convergence, the resulting estimates of ***β***⋆ are equivalent to the maximum likelihood estimators derived from pooled data. This is because, in GLMs, for maximum likelihood estimators, if both the pooled and distributed algorithms are initialized with the same values for 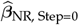 and 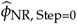, then at each subsequent iteration, the distributed Newton update computed by the CC will be identical to the update obtained in a pooled setting.

For the method to yield consistent estimates, it is not necessary to initialize it with simple averaging estimators. However, using simple averaging estimators as initialization may speed up convergence, particularly in large sample sizes, since these estimators are 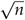 -consistent.

Let 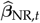 denote the estimator obtained at convergence. Since it is (nearly) equal to the pooled MLE of ***β***^*⋆*^, we can deduce from Appendix B.3.2 that

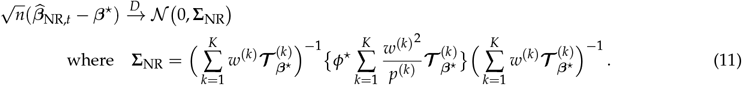

Following the same reasoning used earlier for the single distributed Newton-Raphson update method, we can consistently estimate the variance-covariance matrix as

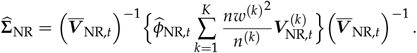

##### Algorithm 2

Distributed Newton-Raphson updatings algorithm

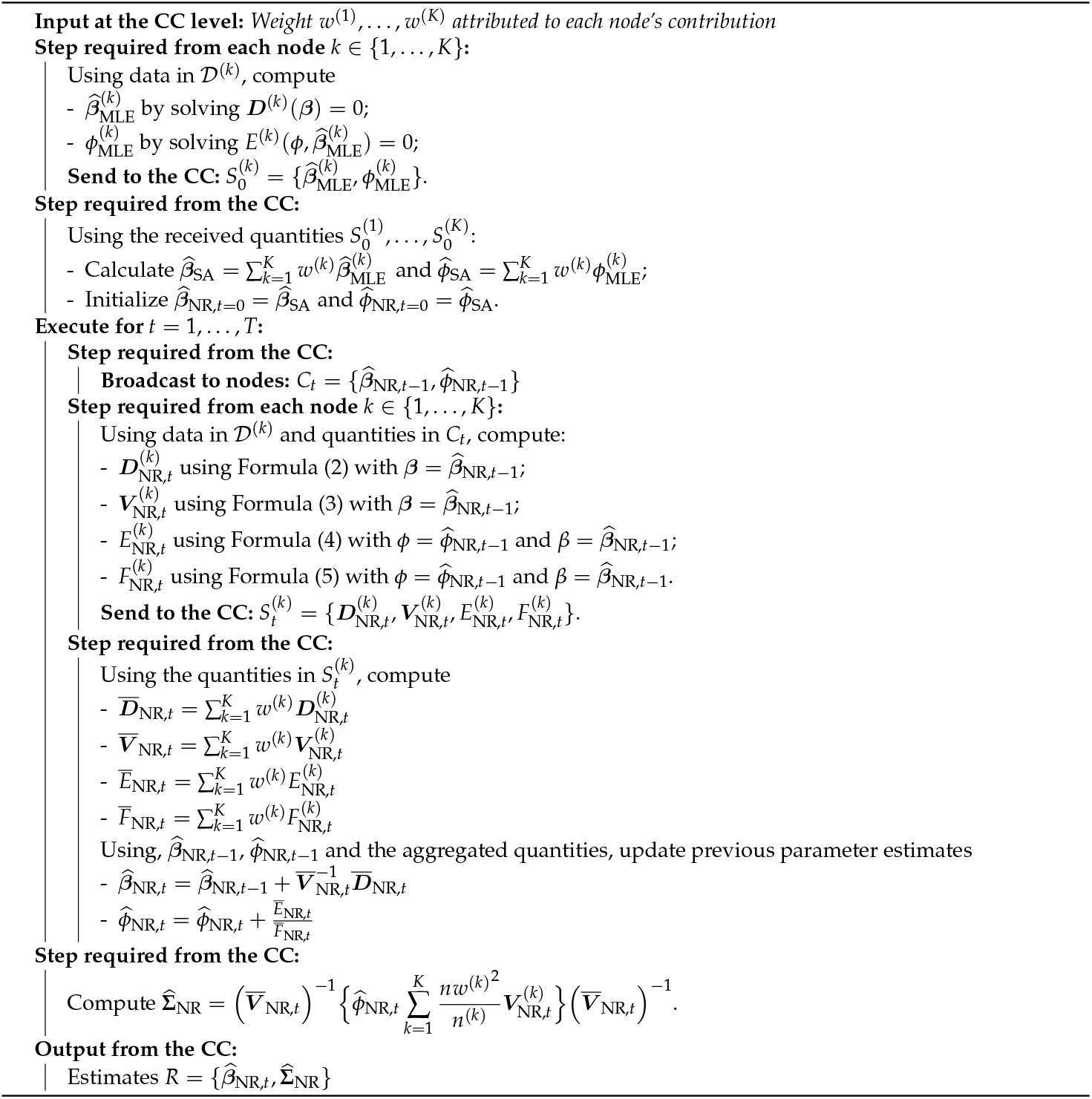

#### 3.3.4 Distributed estimating equation

The distributed estimating equation algorithm follows upon execution of Algorithm 3. First, each node is responsible for computing the MLEs 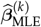 and 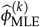 of ***β*\*** and *ϕ*\*, respectively, using its own data. These estimators, along with the hessian matrix 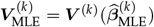 and 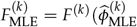, are then sent to the CC. The set

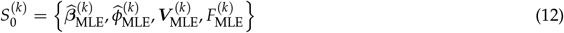

is transmitted to the CC. The CC calculates the weighted average of the Hessians and the 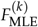 values as follows:

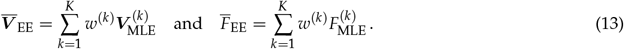

The parameter estimates can then be calculated as

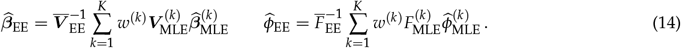

It is shown in Appendix B.3.6 that 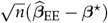 converges in distribution to a centred normal random variable with variance-covariance matrix given by

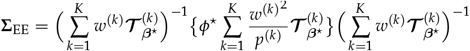

which is equal to **Σ**_NR_. It can be consistently estimated by

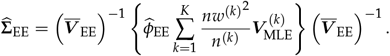

##### Algorithm 3

Distributed estimating equations inference procedure

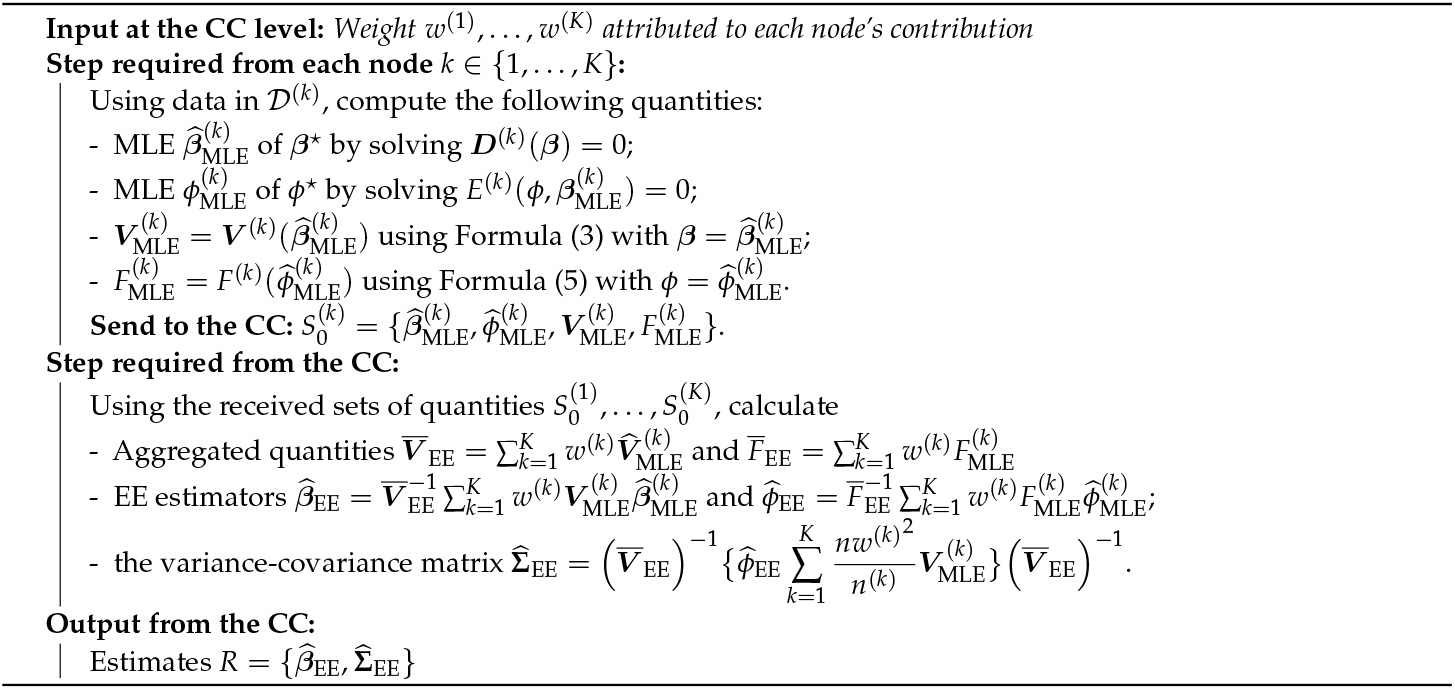

#### 3.3.5 Distributed estimation using a single gradient-enhanced log-likelihood

This method operates through Algorithm 4. First, the necessary information is collected by the CC to compute the initial estimates of ***β*** and *ϕ*, denoted as 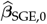 and 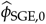. In what follows, we assume these estimates are obtained using the simple averaging estimators calculated through Algorithm 1.

Subsequently, the CC broadcasts 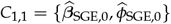 to node *k* ∈ {2, …, *K*}. Each node is then requested to compute and transmit back the following quantities:

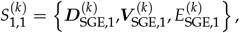

with 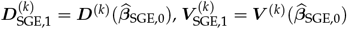 and 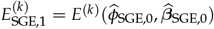.

The CC aggregates the ***D***^(*k*)^’s and the *E*^(*k*)^’s using averaging by calculating

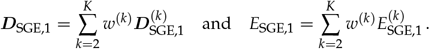

The ***V*** ^(*k*)^’s are momentarily stored and will be used later to compute the estimator for the asymptotic variance-covariance matrix of the final estimator of ***β***^*⋆*^. The quantities

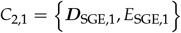

are then sent to node *k* = 1. Node *k* = 1 computes the global average of the ***D***^(*k*)^’s by adding its own counterpart, i.e., it first computes

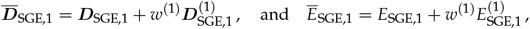

and then solves the surrogate likelihood function. Formally, it finds successively the values 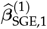 and 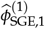 that solve

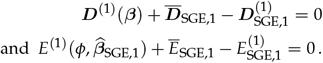

The results are sent back to the CC, along with 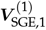, yielding

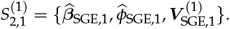

If simple averaging estimators for 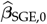 and 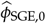 are chosen, then 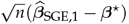 converges in distribution to a mean-zero multivariate normal random variable with variance-covariance matrix given by

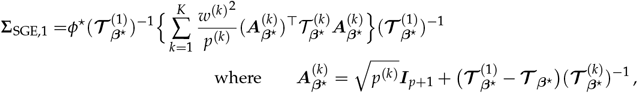

with ***I***_*p*+1_ the *p* + 1 square identity matrix and 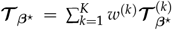. See Appendix B.3.7. The latter can be consistently estimated by

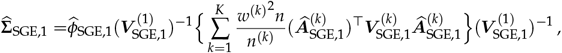

where

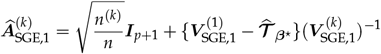

with 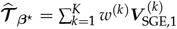.

##### Remark 1.

*In the paper [24], where the method described above was originally proposed, the authors discuss a version in which the latter process is repeated multiple times. Their version assumes that the data is uniformly and randomly split across nodes. Under this assumption, the resulting estimator of* ***β***^*⋆*^ *is asymptotically equivalent to the pooled estimator, regardless of the number of iterations executed. This equivalence occurs because when the predictors’ distribution is the same across nodes and the node sample sizes are equal, then* 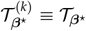 *and p*^(*k*)^ ≡ 1/*K, it follows that* 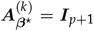 *resulting in the following expression for* Σ_*SGE,1*_:

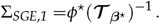

*The variance-covariance matrix above is also the same as that of the simple averaging estimator in the setting of equal sample sizes and even predictor distributions. Consequently, at each iteration, the probability distribution of the resulting estimator remains unchanged. However, in a more general setting where predictor distributions and sample sizes vary across nodes, these cancellations no longer occur. Therefore, in this case, the probability distribution of the obtained estimator changes after each iteration, and tracking these changes falls beyond the scope of objective 3, see Appendix B.3.7. Hence, the current presentation focused on the case where only one iteration is executed*.

##### Algorithm 4

Inference procedure based on the distributed estimation using a single gradient-enhanced log-likelihood method

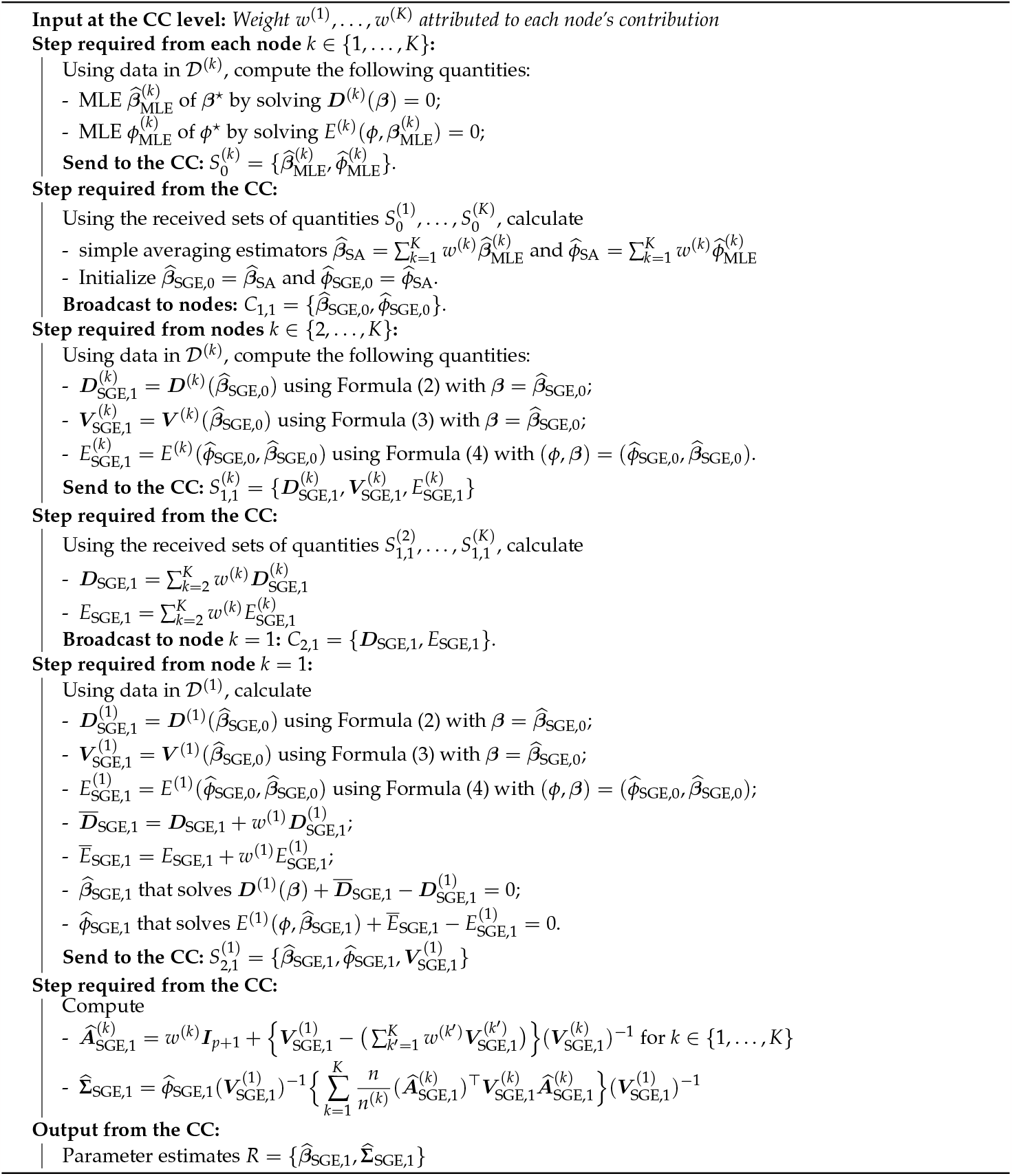

#### 3.3.6 Distributed estimation using multiple gradient-enhanced log-likelihood

This method operates through Algorithm 5. First, the CC collects the necessary information to compute the initial estimates, denoted as 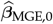 and 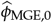. In this case, we assume that these estimates are obtained using the simple averaging estimators calculated through Algorithm 1.

Subsequently, the CC broadcasts 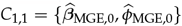 to each node, which is then requested to compute and transmit back the following quantities:

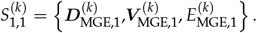

Here, 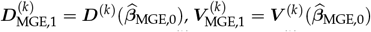 and 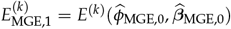.

The CC aggregates the ***D***^(*k*)^’s and the *E*^(*k*)^’s using averaging by calculating

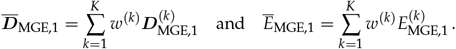

The CC then broadcasts 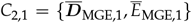 to each node, which are then tasked to solve the surrogate likelihood function. Formally, they find successively the value 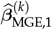 and 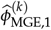 that solves

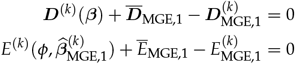

Each node then transmits their set of local surrogate likelihood estimators to the CC:

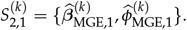

Using the received sets of quantities 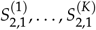, the CC aggregates them through averaging using the following formulas:

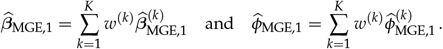

It is shown in Appendix B.3.8 that 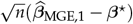 converges in distribution to a multivariate normal random variable with mean 0 and a variance-covariance matrix given by:

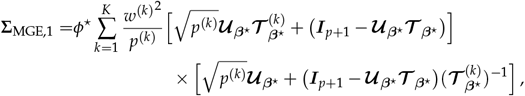

where 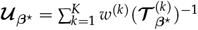. The latter can be consistently estimated with

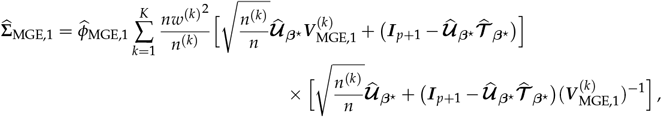

where 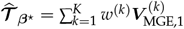 and 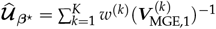.

##### Algorithm 5

Inference procedure based on the distributed estimation using multiple gradient-enhanced log-likelihood method

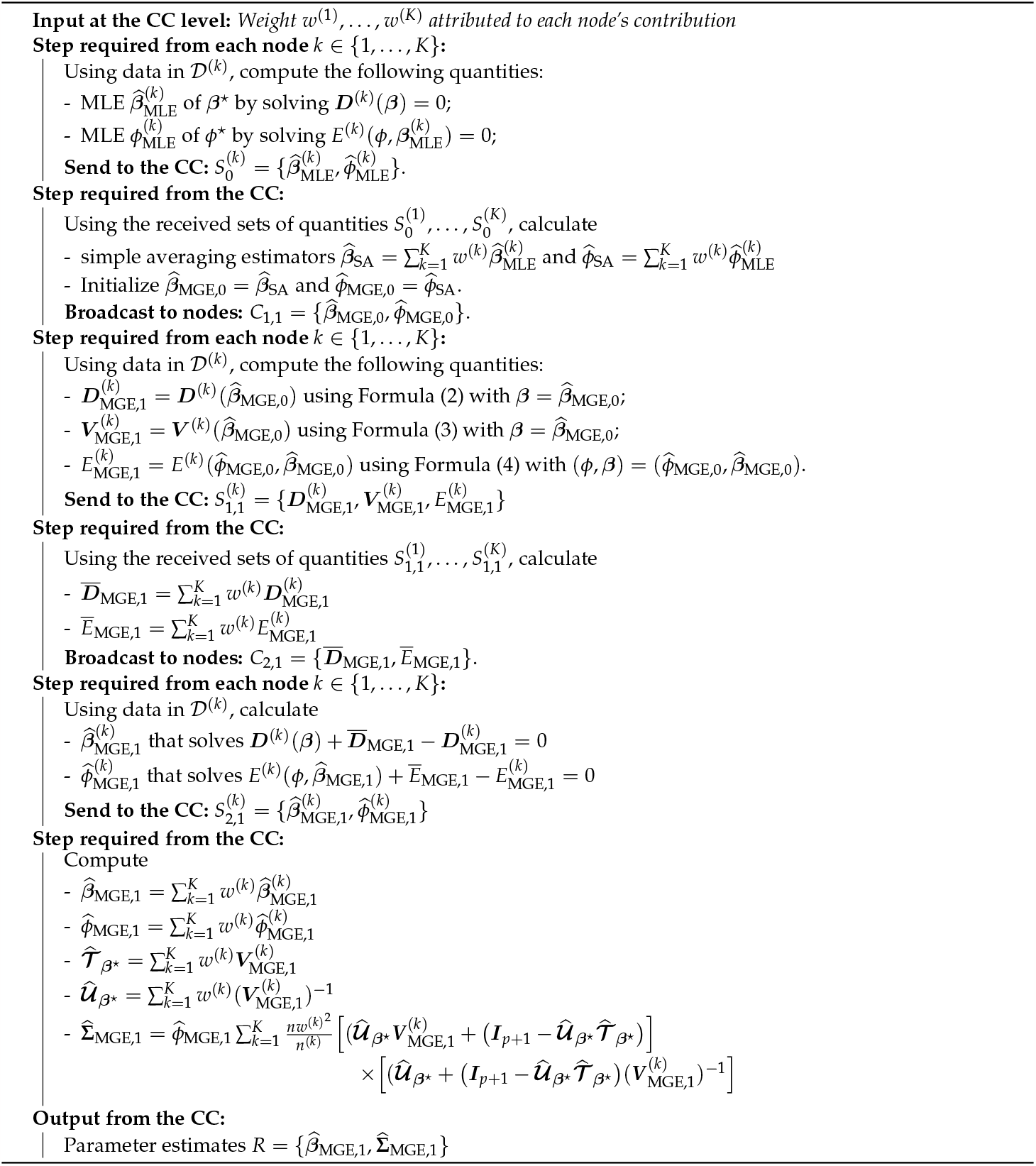

#### 3.3.7 Summary of quantities exchanged for the adapted methods

The following table presents a summary of the quantities exchanged between the nodes and the CC in both directions. Table 2 demonstrates that the quantities involved in exchanges from the nodes to the CC consist of parameter estimates, gradients (***D***^(*k*)^ vectors), Hessians (***V*** ^(*k*)^ matrices), as well as real numbers (*E*^(*k*)^ and *F*^(*k*)^). On the other hand, the quantities shared from the CC to the nodes primarily consist of parameter estimates. Notably, Methods 5 and 6 differentiate themselves by requiring the sharing of aggregated gradient vectors and Hessian matrices as well.

**Table 2:** Quantities shared in each adapted method’s communication workflow.

| Method | Exchanged quantities from nodes to CC |  | Exchanged quantities from CC to nodes |
| --- | --- | --- | --- |
| | $S_0^{(k)}$ | $S_t^{(k)}, t \geq 1$ | $C_t$ |
| 1. Simple averaging | $\hat{\beta}_{\text{MLE}}^{(k)}; \hat{\phi}_{\text{MLE}}^{(k)}; V_{\text{MLE}}^{(k)}$ | - | - |
| 2. Single distributed Newton-Raphson updating | $\hat{\beta}_{\text{MLE}}^{(k)}; \hat{\phi}_{\text{MLE}}^{(k)}$ | $D_{\text{NR},1}^{(k)}; V_{\text{NR},1}^{(k)}; E_{\text{NR},1}^{(k)}; F_{\text{NR},1}^{(k)}$ | $\hat{\beta}_{\text{NR},0}; \hat{\phi}_{\text{NR},0}$ |
| 3. Multiple distributed Newton-Raphson updates (with $T$ Newton-Raphson updates) | $\hat{\beta}_{\text{MLE}}^{(k)}; \hat{\phi}_{\text{MLE}}^{(k)}$ | $D_{\text{NR},t}^{(k)}; V_{\text{NR},t}^{(k)}; E_{\text{NR},t}^{(k)}; F_{\text{NR},t}^{(k)}$ | $\hat{\beta}_{\text{NR},t-1}; \hat{\phi}_{\text{NR},t-1}$ |
| 4. Distributed estimating equations | $\hat{\beta}_{\text{MLE}}^{(k)}; \hat{\phi}_{\text{MLE}}^{(k)}; V_{\text{MLE}}^{(k)}; F_{\text{MLE}}^{(k)}$ | - | - |
| 5. Distributed single gradient-enhanced log-likelihood | $\hat{\beta}_{\text{MLE}}^{(k)}; \hat{\phi}_{\text{MLE}}^{(k)}$ | **Nodes 2 to $K$:**<br>$D_{\text{SGE},1}^{(k)}; V_{\text{SGE},1}^{(k)}; E_{\text{SGE},1}^{(k)}$<br>**Node 1:**<br>$\hat{\beta}_{\text{SGE},1}; \hat{\phi}_{\text{SGE},1}; V_{\text{SGE},1}^{(1)}$ | $\hat{\beta}_{\text{SGE},0}; \hat{\phi}_{\text{SGE},0}$<br>**To node 1 only:**<br>$D_{\text{SGE},1}; E_{\text{SGE},1}$ |
| 6. Distributed multi-gradient-enhanced log-likelihood | $\hat{\beta}_{\text{MLE}}^{(k)}; \hat{\phi}_{\text{MLE}}^{(k)}$ | $D_{\text{MGE},1}^{(k)}; V_{\text{MGE},1}^{(k)}; E_{\text{MGE},1}^{(k)}; \hat{\beta}_{\text{MGE},1}^{(k)}; \hat{\phi}_{\text{MGE},1}^{(k)}$ | $\hat{\beta}_{\text{MGE},0}; \hat{\phi}_{\text{MGE},0}; \bar{D}_{\text{MGE},1}; \bar{E}_{\text{MGE},1}$ |

#### 3.3.8 Comparison of adapted methods

Table 3 compares the main adapted HPSA methods on the quantities shared between the CC and nodes and the operational complexity of the procedures.

**Table 3:**
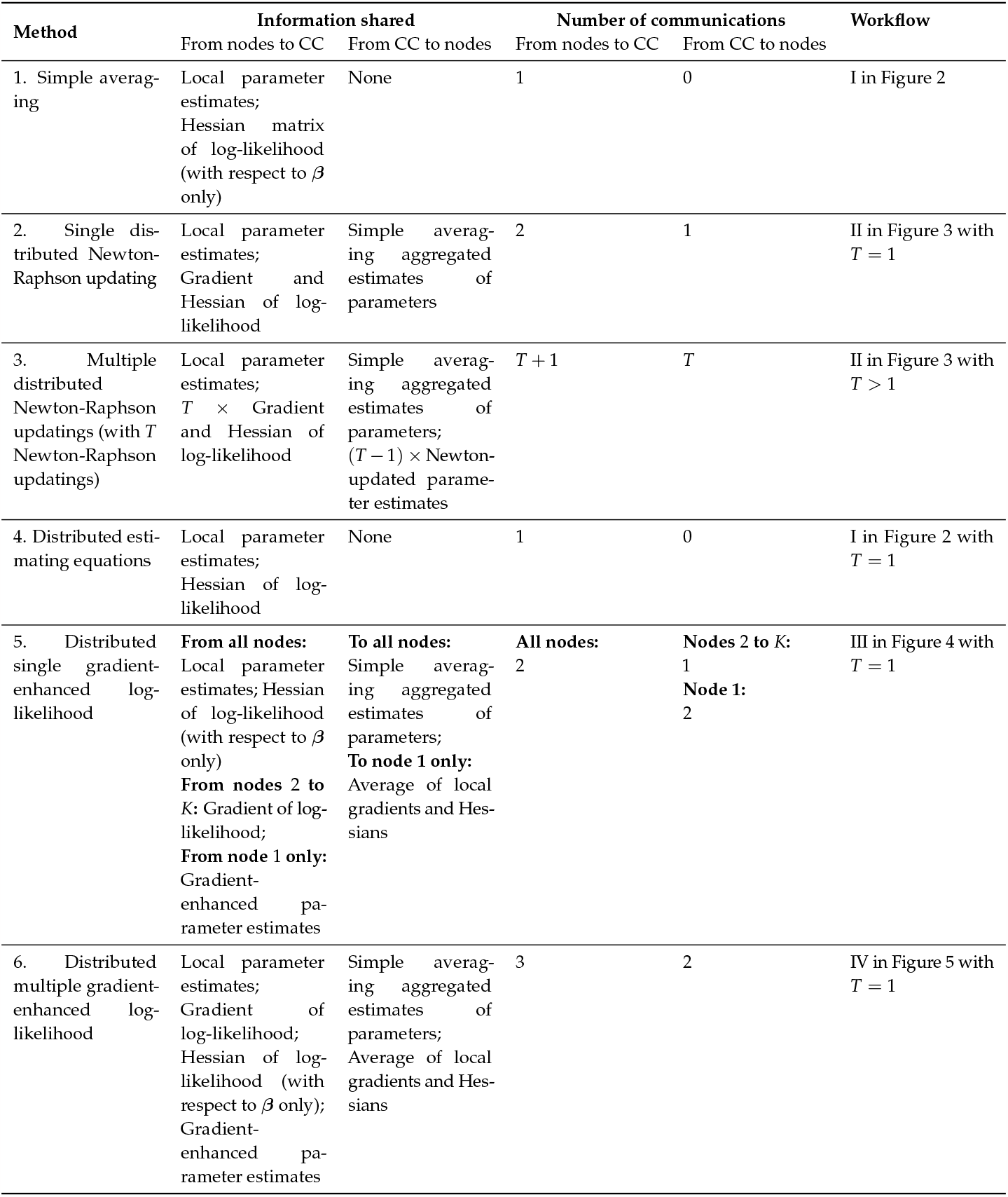
Comparison of adapted methods.

| Method | Information shared |  | Number of communications |  | Workflow |
| --- | --- | --- | --- | --- | --- |
|  | From nodes to CC | From CC to nodes | From nodes to CC | From CC to nodes |  |
| 1. Simple averaging | Local parameter estimates; Hessian matrix of log-likelihood (with respect to $\beta$ only) | None | 1 | 0 | I in Figure 2 |
| 2. Single distributed Newton-Raphson updating | Local parameter estimates; Gradient and Hessian of log-likelihood | Simple averaging aggregated estimates of parameters | 2 | 1 | II in Figure 3 with $T = 1$ |
| 3. Multiple distributed Newton-Raphson updatings (with $T$ Newton-Raphson updatings) | Local parameter estimates; $T \times$ Gradient and Hessian of log-likelihood | Simple averaging aggregated estimates of parameters; $(T - 1) \times$ Newton-updated parameter estimates | $T + 1$ | $T$ | II in Figure 3 with $T > 1$ |
| 4. Distributed estimating equations | Local parameter estimates; Hessian of log-likelihood | None | 1 | 0 | I in Figure 2 with $T = 1$ |
| 5. Distributed single gradient-enhanced log-likelihood | **From all nodes:** Local parameter estimates; Hessian of log-likelihood (with respect to $\beta$ only)<br>**From nodes 2 to $K$:** Gradient of log-likelihood;<br>**From node 1 only:** Gradient-enhanced parameter estimates | **To all nodes:** Simple averaging aggregated estimates of parameters;<br>**To node 1 only:** Average of local gradients and Hessians | **All nodes:** 2 | **Nodes 2 to $K$:** 1<br>**Node 1:** 2 | III in Figure 4 with $T = 1$ |
| 6. Distributed multiple gradient-enhanced log-likelihood | Local parameter estimates; Gradient of log-likelihood; Hessian of log-likelihood (with respect to $\beta$ only); Gradient-enhanced parameter estimates | Simple averaging aggregated estimates of parameters; Average of local gradients and Hessians | 3 | 2 | IV in Figure 5 with $T = 1$ |

**Table 4:**
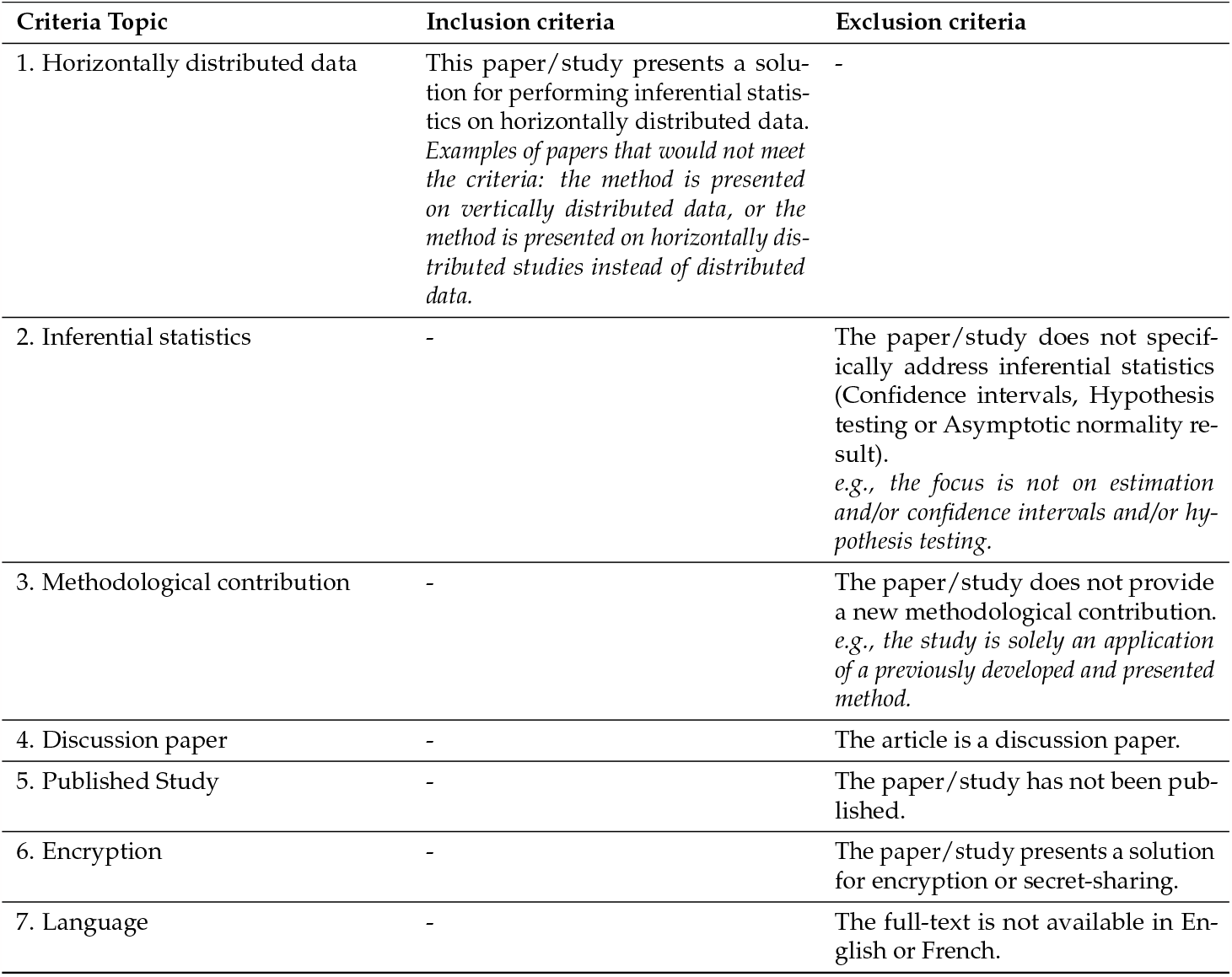
Inclusion and Exclusion criteria.

| Criteria Topic | Inclusion criteria | Exclusion criteria |
| --- | --- | --- |
| 1. Horizontally distributed data | This paper/study presents a solution for performing inferential statistics on horizontally distributed data. *Examples of papers that would not meet the criteria: the method is presented on vertically distributed data, or the method is presented on horizontally distributed studies instead of distributed data.* | - |
| 2. Inferential statistics | - | The paper/study does not specifically address inferential statistics (Confidence intervals, Hypothesis testing or Asymptotic normality result).<br>*e.g., the focus is not on estimation and/or confidence intervals and/or hypothesis testing.* |
| 3. Methodological contribution | - | The paper/study does not provide a new methodological contribution. *e.g., the study is solely an application of a previously developed and presented method.* |
| 4. Discussion paper | - | The article is a discussion paper. |
| 5. Published Study | - | The paper/study has not been published. |
| 6. Encryption | - | The paper/study presents a solution for encryption or secret-sharing. |
| 7. Language | - | The full-text is not available in English or French. |

**Table 5:**
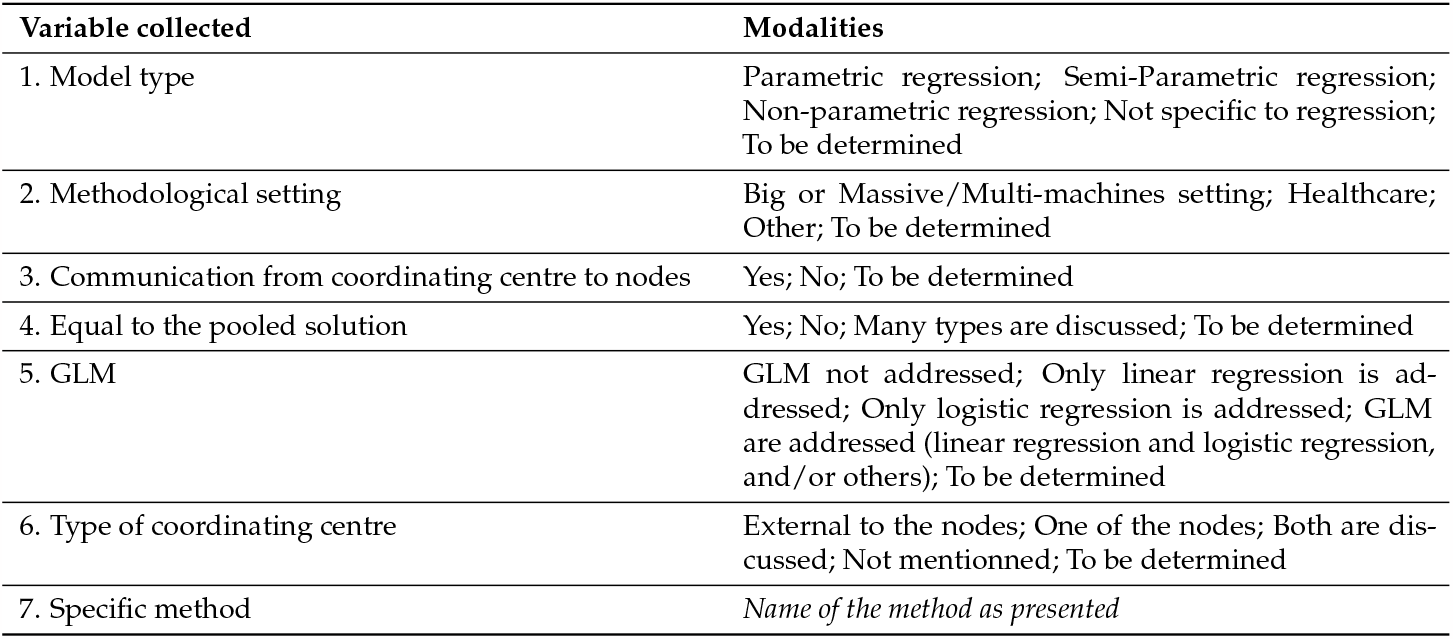
Data extraction.

Methods 1 and 4 require only one communication from the data nodes to the CC and no communication back from the CC to the nodes. These so-called one-shot methods have the lowest operational complexity. Method 4 requires the additional quantity 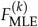 to be transmitted from each node to the CC.

Methods 2 and 3 perform Newton-Raphson updates using some initial estimator as a basis, usually the simple averaging estimator. While Method 2 requires this initial estimator to be 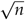 -consistent, if *T* is large enough, any initial value will work for Method 3 (although convergence may be slower). Both methods require ***D***^(*k*)^, ***V*** ^(*k*)^, *E*^(*k*)^ and *F*^(*k*)^ to be evaluated and sent to the CC *T* times, with *T* = 1 for Method 2. Compared to Method 1, Method 2 requires the additional quantities ***D***^(*k*)^, *E*^(*k*)^ and *F*^(*k*)^, and Method 3 further requires these quantities to be evaluated and communicated multiple times.

Method 5 relies on an approximation of the log-likelihood function. It requires an initial estimator, usually the simple averaging estimator. This approach treats node 1 differently, making it solve the surrogate log-likelihood using aggregates from the other nodes and its own data. The CC sends the initial estimator to each node, then requires them to evaluate ***D***^(*k*)^, ***V*** ^(*k*)^ and *E*^(*k*)^ and send the result back to the CC once. It averages the results and then communicates them to node 1, which solves the surrogate log-likelihood and sends its results back to the CC.

Method 6 applies Method 5 to every node, making each node solve the surrogate log-likelihood function with its own data before averaging the resulting local estimators.

## 4 Discussion

### 4.1 Summary of Findings

The first objective (O1) of this study aimed to identify and map the methodological approaches used and developed in the literature regarding HPSA. To achieve this, we conducted a scoping review, which included 41 articles following our protocol. These articles were categorized based on the types of models and communication schemes involved, as presented in Table 1. The analysis revealed that the majority of methods included in the scoping review focused on methodological settings associated with massive data. The communication schemes of these methods were demonstrated through Workflows I, II, III and IV.

The second objective (O2) of this study aimed to describe the approaches that can be employed for basic GLM regression analyses and identify the distributional assumptions they require. To accomplish this, we identified six approaches that could be classified within Workflows I-IV. However, a limitation of these methods is that they assume identical node sample sizes and node covariate distributions. This assumption reduces their suitability in settings commonly encountered in healthcare research, where data collecting nodes are prone to generating different covariate distributions.

The third objective (O3) of this study was to present methods that relaxed these assumptions by adapting the approaches identified in O2 to the unequal sample sizes and non-identical covariate sample distribution setting. Additionally, we compared these methods in terms of the information shared and operational complexity. This involved adapting the quantities and estimators described in the original articles and deriving new asymptotic results with relaxed assumptions. We proposed a unified framework for inference procedures utilizing these methods. The framework encompasses both estimation and the construction of confidence intervals, providing detailed steps for both the data nodes and the CC.

### 4.2 Challenges and Opportunities

Work pertaining to O1 illustrated why it is so challenging for researchers and data custodians alike to find information regarding HPSA. While the HPSA literature is very recent (all included articles were published in 2010 or later), the literature is non-homogeneous, and it has not come to a consensus on nomenclature. No universal terminology exists, and different terms are used in the different fields developing and applying HPSA methods. Many specific methods introduced in applied contexts are special cases of more general methods which may or may not be cited. These characteristics make finding useful and efficient keywords arduous. This required adapting our research strategy.

This difficulty is compounded by the fact that statistical inference is not the main focus of most of the HPSA literature. The majority of published work is in the prediction, learning and optimization contexts. As a result, method assumptions are rarely discussed. This can be a problem when adapting these methods for inference. Furthermore, the methodological setting is often assumed to be in the massive data context where data is randomly distributed between nodes. This allows the authors to make strong assumptions on node sample sizes and covariate distributions which may be unrealistic in the confidential data complex where different data sources. These methods cannot be used directly for inference using confidential health data. While some work remains to be done when the structure of association between the covariate and the outcome is heterogeneous between nodes, we adapted widely used methods for when the distribution of covariate and sample sizes between nodes are not identical.

Table 1 illustrates how the majority of HPSA methods are focused on parametric models. Some work has also been done for semi-parametric and non-parametric regression, and some methods are introduced outside of the regression framework (although they can also be applied to regression). Many methods do not require communication of quantities from the CC to the data nodes: they only require one transmission from the nodes to the CC. Given the lack of awareness around HPSA, starting by implementing lower operational complexity methods while providing useful results offers a promising path.

The methods can be implemented “manually” (e.g. via email exchanges), but platforms enabling semi-automated distributed fittings of statistical models have been proposed in the literature (e.g. [7]). On the other hand, explicit descriptions of their algorithms and the quantities exchanged are not always easily accessible and this complicates the evaluation of the tools by data custodians and researchers.

This is especially important since it is essential to clarify here that operating an HPSA algorithm does NOT ensure confidentiality in and of itself.

For example, it is known that sharing sample moments can compromise confidentiality. It can be shown that a set of *n* observations is uniquely determined by its first *n* sample moments [40]. This could prove problematic for methods that rely on sharing the first few moments of each node’s sample, especially if number of observations is low, as the sample could be partially reconstructed by the CC.

The results presented here contribute to this objective by clarifying the workflows and quantities exchanged by each method. Nevertheless, further analysis of the confidentiality preserved by HPSA methods is needed to fully understand the risk associated with the sharing of summary statistics, especially as more rounds of communication between the CC and data nodes are completed. The framework of differential privacy (DP) has been used to guarantee the preservation of confidentiality in a few HPSA methods, but a wider application of DP to existing and popular methods has yet to be explored.

## Informed consent

Not applicable.

## Data availability

Not applicable.

## Acknowledgments

We would like to thank the GRIIS members who enriched this work via multiple conversations over the last few months and kept us going. We would also like to thank Pr. Kim McGrail for her very insightful comments on this work.

## Conflicts of interest

The authors declare no conflict of interest.

### Abbreviations

CC: Coordinating centre
GLM: Generalized linear model
HPSA: Horizontally Partitioned Statistical Analytics
ICES: Institute for Clinical Evaluative Sciences
LHS: Learning health system
MCHP: Manitoba Centre for Health Policy
MLE: Maximum likelihood estimator
PACS: Picture archiving and communication system

## A Detailed protocol for the scoping review

### A.1 Research question

1. What are the existing methods that allow to conduct statistical inference procedures from a horizontally distributed dataset?
  - *Regarding: Methods for different statistical models; Methods for various settings in terms of information shared; Methods for different needs in terms of precision of estimates*.
2. What are the characteristics of these methods to proceed to a systematic categorisation?
  - *Regarding: Type of algorithm; Settings for nodes and coordinating centre; Capacity to reach exact estimates from data pooling*.

### A.2 Methods

The scoping review will be conducted in accordance with the methodological framework from Levac et al. [26] (based on Arksey and O’Malley [2]).

#### A.2.1 Key-words

The following keywords were identified from the **snowballing literature search**:

- *distributed algorithms* [15]
- *distributed estimation* [21]
- *distributed inference* [24]
- *distributed learning* [31]
- *distributed regression* [46] (not included in the scoping review final selection since no new estimation methods are discussed).
- *federated inference*[57] (not included in the scoping review final selection since the paper was not published when the scoping review search was launched)
- *federated estimation* [50] (not included in the scoping review final selection since the paper was not published when the scoping review search was launched)
- *federated learning* [27] (not included in the scoping study review final selection the paper focuses solely on estimation, i.e. no confidence interval computation strategies or hypothesis testing framework are discussed).
- *privacy-protecting algorithm* [38]
- *privacy-preserving algorithm* [14]
- *aggregated inference* [22] (not included in the scoping review final selection since no new estimation methods are discussed).

The following keywords will be used to add conciseness to the topic of statistical inference, to avoid screening machine-learning specific articles:

- *Statistical inference*
- *Confidence interval*
- *Statistical estimation*
- *Hypothesis tests*
- *Significant coefficient, Significance of parameter*

#### A.2.2 Research strategies

In collaboration with a specialist in documentary research at the Université de Sherbrooke, we have selected the following abstract and citation databases: (1) Medline, (2) Scopus, (3) MathSciNet, and (4) zbMATH. The choice of these databases was motivated by the interdisciplinary nature of the research question, which spans the fields of statistics and health.

To develop comprehensive research strategies, we combined the previously mentioned keywords and worked closely with the documentary research specialist.

##### Limits and restrictions

In order to strike a balance between sensitivity and specificity in our research, given the interdisciplinary nature of the topic involving distributed data and statistical inference, we took several considerations into account.

To ensure sensitivity, we opted for interdisciplinary databases that are known to cover a wide range of relevant literature. These include Medline, Scopus, MathSciNet, and zbMATH. By selecting these databases, we aimed to capture a comprehensive set of articles that encompass both statistical and health-related aspects.

On the other hand, to maintain specificity and avoid retrieving a large number of non-relevant articles, we carefully selected keywords that were targeted and specific to our research question. Instead of relying solely on thesauri and synonym search tools, we focused on the vocabulary commonly used in the literature through an extensive overlook (snowballing) approach, particularly for the concept of distributed data. For the concept of statistical inference, we chose synonyms that specifically capture studies centred around this topic.

Furthermore, to keep the scope of our research manageable and relevant to recent developments, we limited our search to articles published since the year 2000. This restriction is justified by the emergence of distributed data in recent years, driven by advancements in technology and the availability of massive datasets. By setting this threshold, we aimed to capture any early-developed methods and approaches related to our research topic.

Overall, our research strategies were designed to strike a balance between sensitivity and specificity, ensuring that we capture a comprehensive range of relevant articles while minimizing the inclusion of non-relevant ones.

##### Medline search querry

~~~
((AB ((((“Privacy-preserving” OR “Privacy-protecting*” OR “federated” OR “Distributed” OR “aggregated
 “) N1 (“estimation*” OR “algorithm*” OR “inference” OR “analy*” OR “regression*” OR “model*” OR “
 statistic*” OR “learning”))) OR TI ((((“Privacy-preserving” OR “Privacy-protecting*” OR “federated”
 OR “Distributed” OR “aggregated”) N1 (“estimation*” OR “algorithm*” OR “inference” OR “analy*” OR “
 regression*” OR “model*” OR “statistic*” OR “learning”)))) OR SU ((((“Privacy-preserving” OR “
 Privacy-protecting*” OR “federated” OR “Distributed” OR “aggregated”) N1 (“estimation*” OR “algorithm
 *” OR “inference” OR “analy*” OR “regression*” OR “model*” OR “statistic*” OR “learning”))))) AND (
 TX ((“statistical inference” OR “confidence interval*” OR “Statistical Estimat*” OR “hypothesis test
 *” OR “significant coefficient*” OR “significant parameter*”))))
~~~

##### Scopus search querry

~~~
TITLE-ABS-KEY ((“Privacy-preserving” OR “Privacy-protecting*” OR “federated” OR “Distributed” OR “
 aggregated”) W/1 (“estimation*” OR “algorithm*” OR “inference” OR “analy*” OR “regression*” OR “model
 *” OR “statistic*” OR “learning”) AND (“statistical inference” OR “confidence interval*” OR “
 Statistical Estimat*” OR “hypothesis test*” OR “significant coefficient*” OR “significant parameter*”))
~~~

##### MathSciNet search querry

~~~
“Anywhere=(“Privacy-preserving” OR “Privacy-protecting” OR “federated” OR “Distributed” OR “aggregated”)
 AND Anywhere=(“estimation*” OR “algorithm*” OR “inference” OR “analy*” OR “regression*” OR “model*” OR
 “statistic*” OR “learning”) AND Anywhere=(“statistical inference” OR “confidence interval*” OR “
 Statistical Estimat*” OR “hypothesis test*” OR “significant coefficient*” OR “significant parameter*”)
 “
~~~

##### zbMATH search querry

~~~
((ti:(“Privacy-preserving” | “Privacy-protecting” | “federated” | “Distributed” | “aggregated”) \& ti:(“
 estimation*” | “algorithm*” | “inference” | “analy*” | “regression*” | “model*” | “statistic*” | “
 learning”)) | (ut:(“Privacy-preserving” | “Privacy-protecting” | “federated” | “Distributed” | “
 aggregated”) \& ut:(“estimation*” | “algorithm*” | “inference” | “analy*” | “regression*” | “model*” |
 “statistic*” | “learning”))) \& any:(“statistical inference” | “confidence interval*” | “Statistical
 Estimat*” | “hypothesis test*” | “significant coefficient*” | “significant parameter*”)
~~~

##### Grey literature

As one of the exclusion criteria is to exclude all unpublished studies, no research was conducted among grey literature.

#### A.2.3 Selection process

After removing duplicate references, a manual review of the selected references obtained from the databases was conducted to identify relevant articles that address the research question. In this study, a two-stage selection process was employed to ensure a thorough and systematic approach.

To ensure consistency and minimize bias, all reviewers involved in the selection process met before the commencement of the first stage of selection. This initial meeting aimed to establish a shared understanding of the inclusion criteria and research objectives. By aligning their interpretations and definitions of the inclusion criteria, the reviewers ensured a consistent approach throughout the selection process.

During the selection process, there has been a midpoint meeting among the reviewers after the completion of the first stage of selection. This meeting served as an opportunity to discuss any questions, challenges, or uncertainties that may have arisen during the initial selection. By addressing these issues collectively, the reviewers maintained consistency and addressed discrepancies in their evaluations.

Finally, at the end of the second stage of selection, the reviewers had a final meeting. This meeting allowed for a comprehensive discussion of the selected references and ensured that the final set of included articles met the predefined criteria and effectively addressed the research question.

By conducting regular meetings throughout the selection process and discussing the inclusion criteria, the reviewers aimed to maintain consistency, minimize subjectivity, and enhance the reliability of the article selection.

#### A.2.4 Stages of the selection

##### Selection 1: Titles and Abstracts

All titles and abstracts of the references identified through the research strategy were evaluated by a single author (MPD or FCL). Since this step involved a single reviewer, references that were clearly unrelated to the research question or did not meet the inclusion criteria were automatically excluded from further consideration.

The evaluation process conducted by the single author aimed to swiftly discard references that were obviously irrelevant to the research question. This initial screening helped streamline the subsequent stages of the selection process by removing references that did not align with the study’s objectives or criteria.

##### Selection 2: Full text

The full texts of the references selected in the first stage were reviewed by two authors (MPD and FCL). In instances where there were differing opinions between the two initial reviewers, they engaged in discussions to reach a consensus. To ensure impartiality and a final resolution, a third author (JFE) conducted a third review, overseeing the process and making the ultimate decision in cases where disagreements persisted.

##### Additional strategy

The list of references from all the included articles after the selection process was carefully assessed to identify any additional articles that may not have been captured during the initial screening due to specific keywords. This step aimed to ensure a comprehensive approach by exploring the reference lists of the included articles for relevant references that might have been missed in the initial search.

Through this approach, the review aimed to minimize the possibility of excluding relevant studies and to provide a comprehensive and robust synthesis of the available literature on the subject matter.

##### Inclusion criteria

The following criteria were utilized to guide the selection process. Exclusion was considered for a reference if it met at least one of the exclusion criteria, or if it failed to meet at least one of the inclusion criteria.

#### A.2.5 Data-charting

A data-charting form was collaboratively developed to facilitate the extraction of relevant information from the selected studies. The extraction process was conducted manually, with two authors (MPD and FCL) independently extracting data from the first five studies. Subsequently, the authors convened to verify the adequacy of the process and ensure consistency in data extraction. The remaining studies were then divided between the two authors for data extraction.

During the data extraction phase, specific information pertaining to the research questions was identified and recorded. To account for any uncertainties or variables requiring additional review, a “To be determined” modality was included for each extracted variable. This modality serves as a reminder for a second author to review and validate the extracted data, ensuring accuracy and reliability.

## B Mathematical derivations pertaining to Objective 3

### B.1 Notations used in the Appendix

Recall that in the current setting, there are *n* individuals horizontally partitioned across *K* data storage nodes. Each node’s dataset is 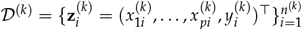, where 1 *≤ k ≤ K* and 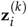 represents measurements on the *i*^th^ individual at node 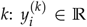 denotes their response variable and 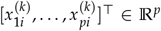 denotes their covariate vector. *n*^(*k*)^ is the total sample size at node *k*. The combined datasets 𝒟 ^(1)^, …, 𝒟 ^(*K*)^ make up the whole dataset without any duplicated individuals such that 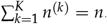.

The current GLM framework assumes that there exists unknown parameters ***β***^*⋆*^ *∈* ℝ ^*p*+1^ *∈* ℝ and *ϕ*^*⋆*^ *>* 0, and known model-specific functions *b, c, g, h* such that with 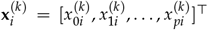 and 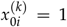, we have 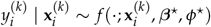, where for any ***β*** = [*β*_0_, *β*_1_, …, *β*_*p*_]^*⊤*^ *∈* ℝ ^*p*+1^ and *ϕ*,

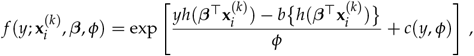

where *b* is such that 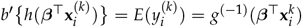, with *b*^*′*^ (*x*) = *∂b*(*x*)/*∂x*.

We also recall the definition of ***D***^(*k*)^(***β***) *∈* ℝ ^*p*+1^ at page 13:

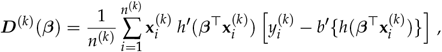

as well as the one of ***V*** ^(*k*)^(***β***) at page 13:

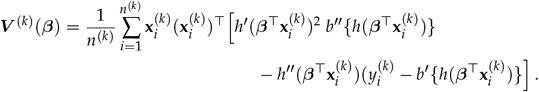

Finally, let us reiterate the definitions of *E*^(*k*)^ in equation (4) and *F*^(*k*)^ in equation (5), which are expressed as follows:

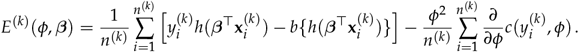

and

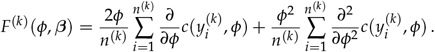

### B.2 General estimation in a pooled centralized setting

The likelihood of the full dataset 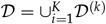, in a setting where the likelihood contribution of each node would be given by the set of weights 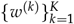, is given by

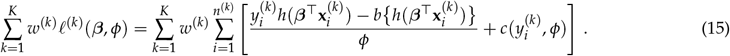

Pooled maximum likelihood estimates of ***β***^*⋆*^ and *ϕ*^*⋆*^ are found by calculating a set of values 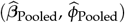 that maximizes (15). This is usually done in two steps. In a first step, equating the gradient with respect to the ***β*** parameters to 0 yields a set of equations that are independent of *ϕ* which, in our framework, are given by

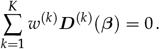

As *g*^(*−*1)^ is often non-linear, iterative methods are necessary to solve the latter equations. When a solution exists and is unique (this is the case under general conditions [53]), the resulting estimator 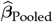 is called the *maximum likelihood estimator*.

In a second step, using 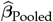, a maximum likelihood estimator of *ϕ*^*⋆*^ can be obtained by solving

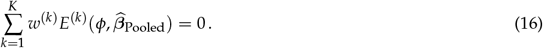

The above equations can be further reduced when 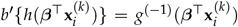, which happens when *g* is canonical, since in this case, *h*(*x*) *≡ x*.

When *ϕ*^*⋆*^ is unknown, it can be estimated by differentiating the log-likelihood at 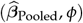 with respect to *ϕ* and equating it to 0. Indeed, since the likelihood equations of ***β*** do not involve *ϕ*, it always holds that

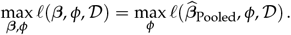

Proceeding in this way yields the following equation for a maximum likelihood estimator of *ϕ* to satisfy:

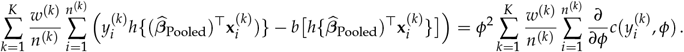

### B.3 Calculations related to unequal sample sizes and uneven between-nodes covariate distributions

The theoretical validity of each algorithm presented in section 3.3 relies on two main components:

1. An asymptotic normality result for the estimator of the ***β*** parameters involved;
2. The consistency, i.e., convergence in probability to the true value, of the estimator for the asymptotic variance-covariance matrix involved in the aforementioned asymptotic normality result. This, in turn, depends on the consistency of the estimator of *ϕ* when the latter is unknown.

Since the current paper is already quite extensive, we will provide theoretical arguments for the asymptotic normality result only, as it is arguably the most interesting from a theoretical perspective. The proof of consistency of the variance-covariance matrix is a lengthy and technical exercise that can be accomplished using our arguments in combination with standard M-estimation theorems, which can be found, for example, in [48], chapter 5.

#### B.3.1 Conditions used to establish asymptotic normality results

The following conditions will be used. For *ℓ ∈* {0, 1, 2, 3}, let

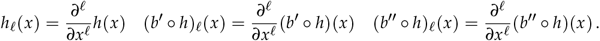

Also, in what follows, for any vector ***a*** ∈ ℝ ^*p*+1^, one defines ∥***a***∥_∞_ = max_1*≤j≤p*+1_ |[***a***]_*j*_| and 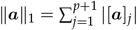.

##### Conditions C

(C1) For *k* ∈ {1, …, *K*}, *n*^(*k*)^/ →*n p*^(*k*)^ *>* 0 as *n* → ∞, and *K* ≥ 2 is finite;

(C2) *b* and *h* are three times continuously differentiable;

(C3) For 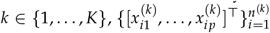 is a set of i.i.d. random vectors with finite sixth marginal moments, i.e., 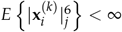, and 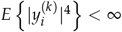. Further, 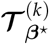 is positive definite, where

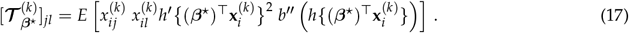

(C4) The ***β***-parameter space Θ *⊂* ℝ^*p*+1^ considered for the search of ***β***^*⋆*^ is compact, and ***β***^*⋆*^ lies in the interior of Θ. Further, one has *E*{***D***^(*k*)^(***β***)} = 0 if and only if ***β*** = ***β***^*⋆*^.

(C5) For 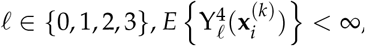, where ϒ_*ℓ*_(**x**) = sup |*h*_*ℓ*_(***β***^*⊤*^**x**)|. Moreover, for 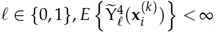 and 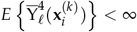, where 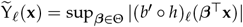 and 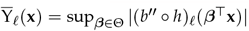.

Assumption (C1) states that each data node has a non-negligible proportion of the data. Assumption (C2) imposes a smoothness condition on the known quantities involved in the definition of the GLM, enabling the use of standard theoretical arguments to derive the asymptotic normality of the estimated coefficients. It is not restrictive. The assumption (C3) that the within-node predictor distribution is the same across all individuals is made to simplify the arguments and to make them more concise. It could be relaxed in various ways, for example, by assuming equal first and second-order moments of relevant quantities instead of the entire distribution.

The compactness of Θ in Condition (C4) is used to establish that ***D***^(*k*)^(***β***) and ***V*** ^(*k*)^(***β***) are uniformly consistent across all possible values for ***β***^*⋆*^, which is a commonly used assumption in maximum likelihood estimation. The identification condition ensures that ***β***^*⋆*^ is the unique value that maximizes the expectation of the node-specific likelihood.

Assumption (C5) is a technical requirement to establish a uniform consistency result for ***D***^(*k*)^(***β***) and ***V*** ^(*k*)^(***β***). It is satisfied when the first, second, and third-order derivatives of *h* and *b* are bounded, as long as 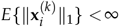. More generally, it imposes a condition on the tails of the distribution of the 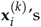. For example, in Poisson regression, where *h*(*x*) = *x* and *b′* (*x*) = *e*^*x*^, this assumption is satisfied if 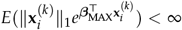, where 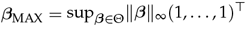 This condition holds, for example, when the 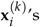 s are normally distributed or have compact support.

#### B.3.2 Theory for the pooled centralized setting estimator

Proceeding as in the proof of Lemma 5 one can show that 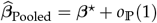. From there, one has, in view of Lemma 7, that

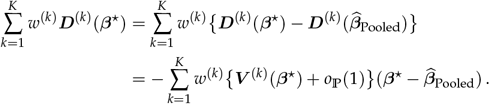

Since 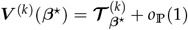 (see Lemma 7), and as ***D***^(*k*)^ is *O*_ℙ_ (*n*^*−*1/2^) (see Lemma 2), one obtains that

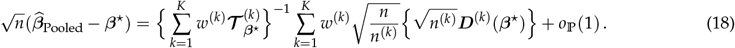

Lemma 2 implies 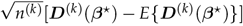 converges in distribution to a centred normal random variable with covariance matrix 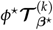 for each 1 *≤ k ≤ K*. Since the 𝒟^(*k*)^’s are mutually independent, as *K* is finite, and because *n*/*n*^(*k*)^ *→* 1/*p*^(*k*)^ as *n →* ∞, then, in view of the above equation, Slutsky’s theorem ensures that

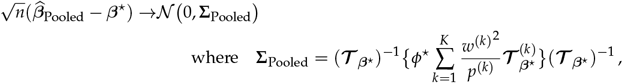

with 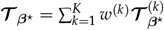.

#### B.3.3 Theory for the adapted simple averaging estimator

Since 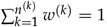, then, using the definition of 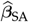, one has

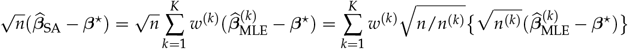

By Lemma 6, under Conditions (C1) to (C5), it holds as *n →* ∞ that, for all 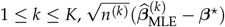 converges in distribution to a centred normal random variable with variance-covariance matrix given by 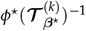. Since the *𝒟* ^(*k*)^’s are mutually independent, as *K* is finite, and because *n*/*n*^(*k*)^ *→* 1/*p*^(*k*)^ as *n →* ∞, then, in view of the above equation, Slutsky’s theorem ensures that

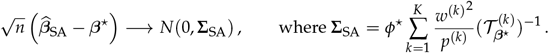

#### B.3.4 Theory for the adapted single distributed Newton-Raphson updating estimator

Let 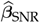 denote the single distributed Newton-Raphson updating estimator 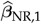 of ***β***^*⋆*^. One has from (10) that

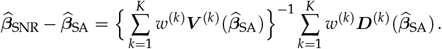

Since 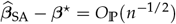,

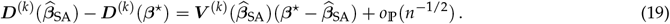

Hence,

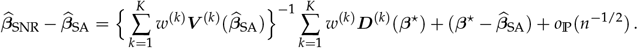

One concludes by re-arranging terms in the preceding equation that

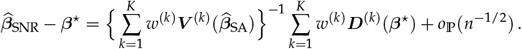

Since Lemma 7 ensures the relationship 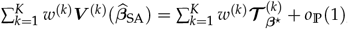, the right-hand side of the last equation is asymptotically equivalent to the right-hand side of (18). Hence, one concludes that

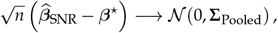

where **Σ**_Pooled_ is as above.

#### B.3.5 Theory for the adapted multiple distributed Newton-Raphson updating estimator

Let 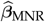 denote the multiple distributed Newton-Raphson updatings estimator. When iterations are conducted until convergence, the obtained estimator of ***β***^*⋆*^ is equal to 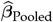. Hence,

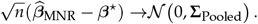

#### B.3.6 Theory for the distributed estimating equations estimator

Let 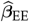 denote the obtained distributed estimating equations estimator. Since it has been established above that 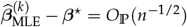 one obtains from a multivariate Taylor expansion that it holds uniformly in *k ∈* {1, …, *K*} and as *n →* ∞ that

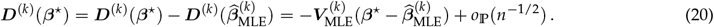

Recalling the definitions of 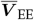 and 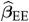 from (13) and (14) we hence have

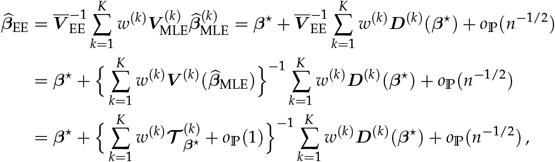

where, to obtain the last line, we used the fact that Lemma 7 ensures 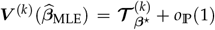. Rearranging terms and considering 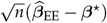, the corresponding right-hand side is then asymptotically equivalent to the right-hand side of (18), and one concludes

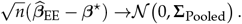

#### B.3.7 Theory for the distributed estimation using a single gradient-enhanced log-likelihood

Let 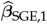 to denote the surrogate likelihood estimator computed at node *k* = 1, and recall that 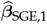 satisfies

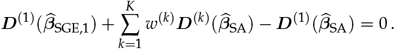

As 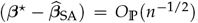, and since Lemma 7 guarantees that 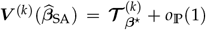, one has from Equation (19) that it holds for each *k ∈* {1, …, *K*} that 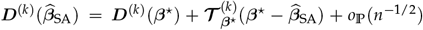.

Hence,

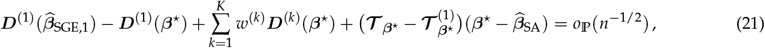

where one recalls that 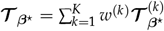.

Next, proceeding as in the proof of Lemma 5 one can show that 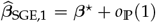. By Lemma 7, the latter result ensures that 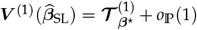. In view of this result, combining the multivariate Taylor’s theorem, the equality *∇*_***β***_ ***D***^(*k*)^(***β***) = *−****V***^(*k*)^ (***β***) and the fact that 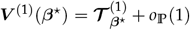 yields the relationship 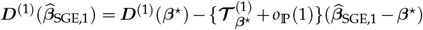 and therefore 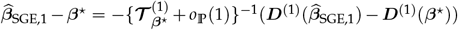. Moreover, in view of Lemma 6 one has

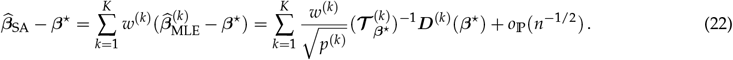

Denoting by ***I***_*p*+1_ the *p* + 1 square identity matrix, one obtains by combining the derived expression for 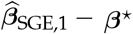 with (21) and (22) that

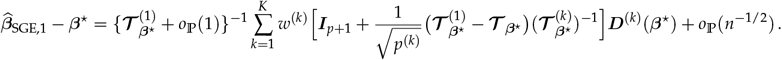

Since the ***D***^(*k*)^’s are *O*_P_(*n*^*−*1/2^) (see Lemma 2), one deduces that

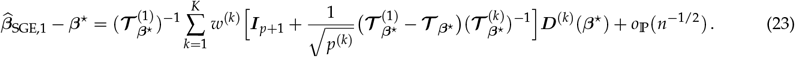

Therefore, 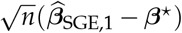 converges in distribution to a mean 0 multivariate normal random variable with variance-covariance matrix given by

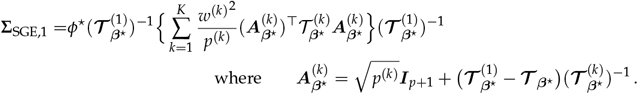

If two iterations are executed, one first uses the fact that

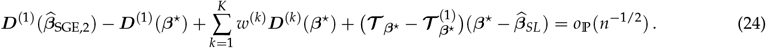

From the last equation, an application of the multivariate Taylor expansion combined with Lemma 2 and Lemma 7 ensures 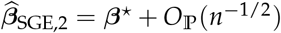. Hence, one obtains that

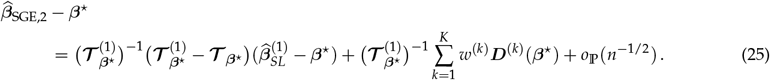

With 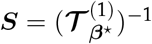 and 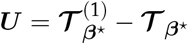, the last equation expresses as

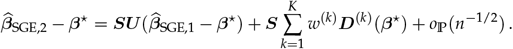

Since from (23) one has

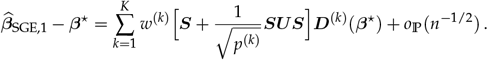

it follows that

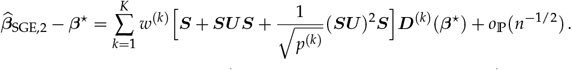

Hence, in general, the asymptotic distribution of 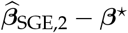 does not match that of 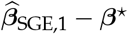 in (23).

#### B.3.8 Theory for the distributed estimation using multiple gradient-enhanced log-likelihoods

Let 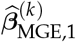 to denote the surrogate likelihood estimator computed at node *k*. Proceeding as we did in the last section to derive (23), one can show that it holds for all *k ∈* {1, …, *K*} that as *n →* ∞,

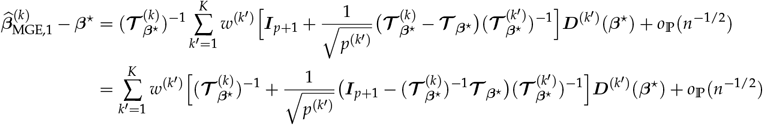

Therefore, letting 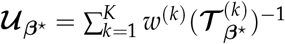, one obtains that

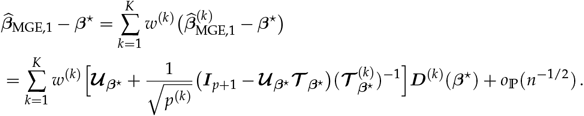

Therefore, 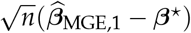 converges in distribution to a mean 0 multivariate normal random variable with variance-covariance matrix given by

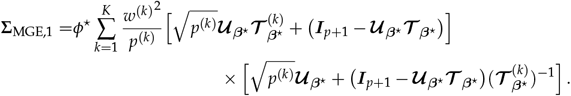

### B.4 Auxiliary results

The following lemma transfers the conditions on the marginal moments imposed in (C3) into a condition on 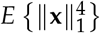 that is used in the proof of Lemma 2.

#### Lemma 1.

*Denote by* **x** *a p* + 1 *dimensional random vector such that* 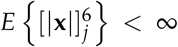 *for all* 1 *≤ j ≤ p* + 1. *Then* 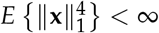.

*Proof*. Note first that for a multiindex ***α*** ∈ ℕ^*p*+1^ such that ∥***α***∥ _1_ = 4 we have essentially 5 possibilities for ***α***: There is one non-zero element 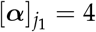 at position *j*_1_, there are two non-zero elements 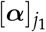 and 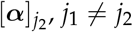, in ***α*** where we either have 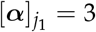 and 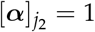 or 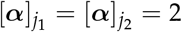, there are three non-zero elements 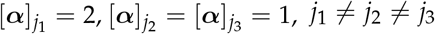, in ***α***, and lastly there are four non-zero elements 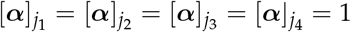 for *j*_1_≠*j*_2_≠*j*_3_≠*j*_4_. Concerning the expectation of ∣ **x ∣** ^***α***^ for a random vector **x** we have by applying the (generalized) Hölder inequality for the five cases that

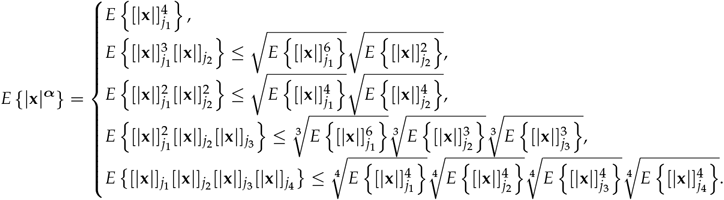

Given that 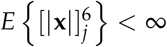 implies also 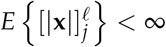 for 1 *≤ ℓ ≤* 5, we see that *E* {|**x**|^***α***^} *<* ∞ for every multiindex ***α*** with ∥***α***∥_1_ = 4. Applying the multinomial theorem to 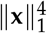 now shows that

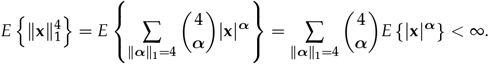

#### Lemma 2.

*Under Conditions (C2)–(C5), it holds that*

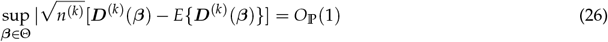

*for all k ∈* {1, …, *K*}.

*Proof*. Let 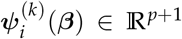 such that 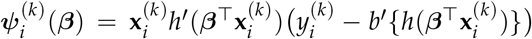. In this notation we have 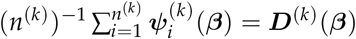. For any ***β***_1_, ***β***_2_ *∈* Θ one has

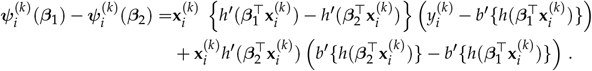

Hence,

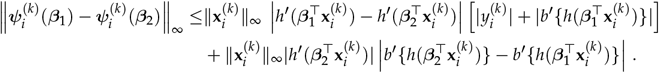

Since by Condition (C2) *h*^*′*^ is differentiable, then, recalling the definition of ϒ_*ℓ*_ in Condition (C5), one deduces from the mean-value theorem and the dual version of the Cauchy–Schwarz inequality |**y**^⊤^ **x**| ≤ ∥**y**∥_∞_∥**x**∥_1_ that

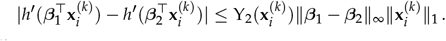

Recalling the definition of 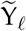 in Condition (C5) one similarly has

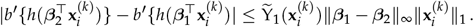

As one also has 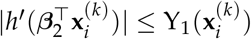 and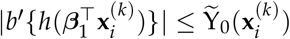, the above equations imply with 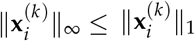 that

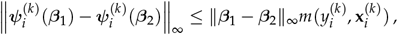

where we set

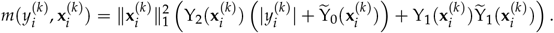

Using first the Hölder and then twice the Minkowski (triangle) inequality we now have

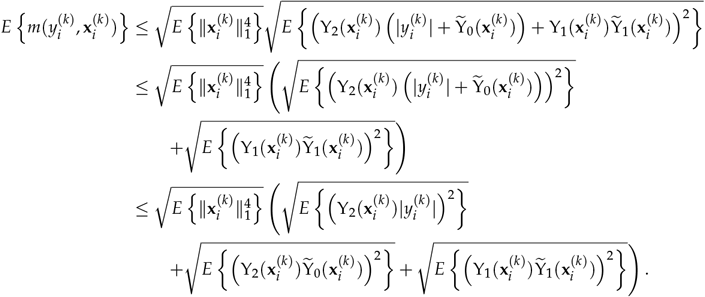

Concerning the individual terms in the parentheses we have by the (generalized) Hölder’s inequality with 1/2 = 1/4 + 1/4 that

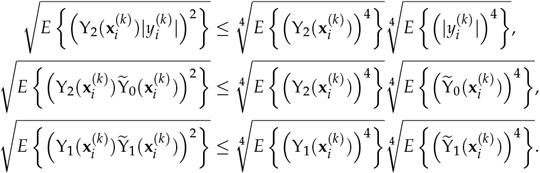

Given these estimates we then have

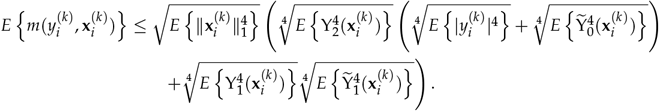

In view of the last equation, Condition (C5) in combination with Condition (C3) and Lemma 1 ensures 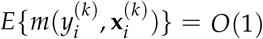. Given that the arguments so far are build on the ∥ · ∥ _∞_ norm, conditions (C3) and (C4) now show that each component

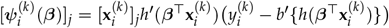

of 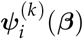 has the property

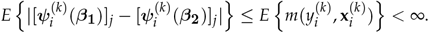

This allows to apply Theorem 19.5 in [48] (see example 19.7) to conclude that each component in bounded in probability. Combining this with [49, Lemma 1.4.3] shows that the same is true when considering all components in 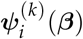 simultaneously. This finally shows that (26) holds.

#### Lemma 3.

*Under Conditions (C2)–(C5), it holds as n →* ∞ *that* 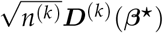 *converges in distribution to a centred normal random variable with covariance matrix*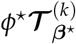.

*Proof*. To prove the Lemma we use the Cramèr-Wold device. That is, we show that, for any constant **a** ∈ ℝ ^*p* +1^, the random variable 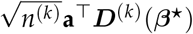 converges in distribution to a centred normal random variable, with variance 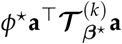. To do this, first note that as 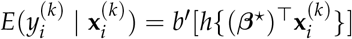, we have *E*{**a**^*⊤*^***D***^(*k*)^(***β***^*⋆*^)} = 0 and

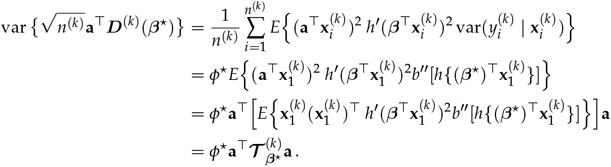

To obtain the second line, we used the fact that var 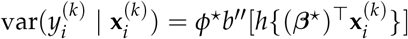 and the assumption that the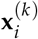 ‘s are i.i.d. for a given *k*. For the third line, we used the equality 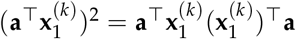.

As the 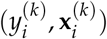 ’s are i.i.d. random variables, it follows 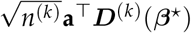 is itself a sum of i.i.d. random variables, with mean 0 and finite (constant) variance. Therefore, an application of the Lindeberg-Lévy central limit theorem ensures that 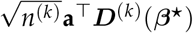 converges in law to a centred normal distribution with variance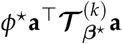.

An application of Cramér-Wold theorem concludes the proof of the Lemma.

The Lemma below can be proven using similar arguments, so their proofs are omitted.

#### Lemma 4.

*Under Conditions (C1) to (C5), it holds as n →* ∞ *that*

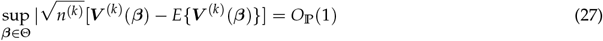

*for all k* ∈ {1, …, *K*}.

The next lemma establishes the consistency of 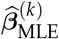.

#### Lemma 5.

*Under Conditions (C1) to (C5), it holds as n →* ∞ *that* 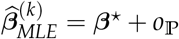(1) *for all* 1 *≤ k ≤ K*.

*Proof*. To prove the Lemma, the goal is the apply Theorem 5.9 in [48] with *θ ≡* ***β***, Ψ_*n*_ *≡* ***D***^(*k*)^ and Ψ *≡ E****D***^(*k*)^. To do this, it is required to verify that (1) it holds as *n →* ∞ that

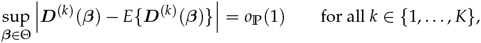

and (2) that for every *ϵ >* 0,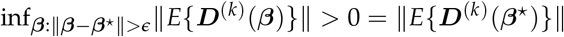.

That (1) holds follows from the fact that under the Lemma’s condition, Lemma 2 applies. That (2) holds follows from the fact that under Condition (C2) the mapping ***β*** *→ E*{***D***^(*k*)^(***β***)} is continuous, and that under Condition (C4) one has *E*{***D***^(k)^ (***β***)} = 0 if and only if ***β*** = ***β***^⋆^. Hence, Theorem 5.9 applies, thereby ensuring that 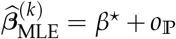 (1).

The fact that it holds for all 1*≤ k ≤ K* follows from the fact that under Condition (C1) *K* is finite.

The last three Lemmas ensure the following result.

#### Lemma 6.

*Under Conditions (C1) to (C5), it holds as n →* ∞ *that, for all* 1 *≤ k ≤ K*,

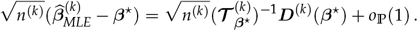

*Consequently*, 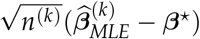 *converges in distribution to a centred normal random variable with variance-covariance matrix given by* 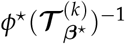.

*Proof*. See Theorem 5.21 in [48].

#### Lemma 7.

*Under Conditions (C1) to (C5), it holds as n →* ∞ *that, for all* 1 *≤ k ≤ K, and any* 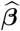 *such that* 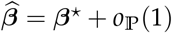,

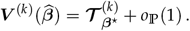

*Proof*. Let Ψ(***β***) = *E*{***V*** ^(*k*)^(***β***)}. Lemma 4 implies that

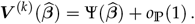

Since it is assumed that 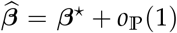 and as 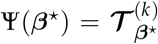, the result follows from the continuous mapping theorem.

